# Oxylipin profiling identifies a mechanistic signature of metabolic syndrome: results from two independent cohorts

**DOI:** 10.1101/2022.03.04.22271895

**Authors:** Céline Dalle, Jérémy Tournayre, Malwina Mainka, Alicja Basiak-Rasała, Mélanie Pétéra, Sophie Lefèvre-Arbogast, Jessica Dalloux-Chioccioli, Mélanie Deschasaux-Tanguy, Lucie Lécuyer, Emmanuelle Kesse-Guyot, Léopold Fezeu, Serge Hercberg, Pilar Galan, Cécilia Samieri, Katarzyna Zatońska, Philip C. Calder, Mads Fiil Hjorth, Arne Astrup, André Mazur, Justine Bertrand-Michel, Nils H. Schebb, Andrzej Szuba, Mathilde Touvier, John W. Newman, Cécile Gladine

**Affiliations:** Université Clermont Auvergne, INRAE, UNH, Clermont-Ferrand, France; Chair of Food Chemistry, Faculty of Mathematics and Natural Sciences, University of Wuppertal, Wuppertal, Germany; Department of Social Medicine, Wroclaw Medical University, Wroclaw, Poland; Université Clermont Auvergne, INRAE, UNH, Plateforme d’Exploration du Métabolisme, MetaboHUB Clermont, Clermont-Ferrand, France; Université de Bordeaux, INSERM, Bordeaux Population Health Research Center, UMR 1219, Bordeaux, France; MetaToul, MetaboHUB, Inserm/UPS UMR 1048 - I2MC, Institut des Maladies Métaboliques et Cardiovasculaires, Toulouse, France; Nutritional Epidemiology Research Team (EREN), Sorbonne Paris Nord University, INSERM U1153, INRAE U1125, Cnam, Epidemiology and Statistics Research Center-University of Paris (CRESS), Bobigny, France; School of Human Development and Health, Faculty of Medicine, University of Southampton, Southampton, United Kingdom; NIHR Southampton Biomedical Research Centre, University Hospital Southampton NHS Foundation Trust and University of Southampton, Southampton, United Kingdom; Department of Nutrition, Exercise and Sports, University of Copenhagen, Copenhagen, Denmark; Department of Angiology, Hypertension and Diabetology; Wroclaw Medical University, Wroclaw, Poland; Obesity and Metabolism Research Unit, United States Department of Agriculture, Agricultural Research Service, Western Human Nutrition Research Center, Davis, CA, USA; University of California Davis Genome Center, University of California, Davis, CA, USA; Department of Nutrition, University of California, Davis, CA, USA

**Keywords:** Oxylipins, Lipidomics, Metabolic syndrome, Biomarker, Validation.

## Abstract

**Background:** Metabolic syndrome (MetS) is a complex condition encompassing a constellation of cardiometabolic abnormalities. Integratively phenotyping the molecular pathways involved in MetS would help to deeply characterize its pathophysiology and to better stratify the risk of cardiometabolic diseases. Oxylipins are a superfamilly of lipid mediators regulating most biological processes involved in cardiometabolic health.

**Methods:** A high-throughput validated mass spectrometry method allowing the quantitative profiling of over 130 oxylipins was applied to identify and validate the oxylipin signature of MetS in two independent case/control studies involving 476 participants.

**Results:** We have uncovered and validated an oxylipin signature of MetS (coined OxyScore) including 23 oxylipins and having high performances of classification and replicability (cross-validated AUC_ROC_ of 89%, 95% CI: 85%-93% and 78%, 95% CI: 72%-85% in the Discovery and Replication studies, respectively). Correlation analysis and comparison with a classification model incorporating both the oxylipins and the MetS criteria showed that the oxylipin signature brings consistent and complementary information supporting its clinical utility. Moreover, the OxyScore provides a unique mechanistic signature of MetS regarding the activation and/or negative feedback regulation of crucial molecular pathways that may help identify patients at higher risk of cardiometabolic diseases.

**Conclusion:** Oxylipin profiling identifies a mechanistic signature of metabolic syndrome that may help to enhance MetS phenotyping and ultimately to better predict the risk of cardiometabolic diseases *via* a better patient stratification.

## 1. Introduction

Metabolic syndrome (MetS) is a serious global public health concern. It is reaching epidemic proportions with a global prevalence estimated to be about one quarter of the world population (1). It is a progressive and heterogeneous condition encompassing a constellation of cardiometabolic abnormalities including central obesity, elevated blood pressure, hypertriglyceridemia, low high density lipoprotein cholesterol (HDLc) and dysglycemia (2) that, when associated, culminate in a 5-fold and 3-fold increased risk of type 2 diabetes mellitus and cardiovascular diseases, respectively (3). Mechanisms underlying MetS are complex and incompletely understood but there is epidemiological evidence showing strong associations with inflammation, oxidative stress, alteration of insulin sensitivity and thrombosis as well as endothelial, renal and hepatic dysfunctions (4).

From a clinical perspective, it is crucial to provide a more complete view of the molecular pathways involved in the onset and development of MetS to better understand, stratify, and ultimately prevent the risk of cardiometabolic diseases (5). In this field, metabolomics and lipidomics are powerful tools that can capture the complexity of MetS and inform on underlying mechanisms at the metabolite level (6). Untargeted data-driven metabolomics has proven its utility to comprehensively characterize the metabolic changes observed in MetS. However, the coverage of untargeted assays is often limited to the most abundant metabolites (such as building blocks of cell membrane and “fuel” metabolites involved in energy production and storage) and does not inform on signaling metabolites that are present at lower concentrations (7). Moreover, the partial mapping of the metabolome and annotation uncertainties limit biological interpretation (8). To face this limitation while keeping a systemic approach, we have targeted a specific class of lipid mediators called oxylipins (including eicosanoids) arising from the oxygenation of polyunsaturated fatty acids (PUFAs) through the coordinated action of over 50 unique and cell-specific enzymes (9). Oxylipins are both biomarkers and mediators of oxidative stress and inflammation, two underlying processes in the pathogenesis of MetS (10, 11). Moreover, oxylipins are involved in the regulation of a vast array of biological processes related to cardiometabolic health including blood clotting, endothelial permeability, blood pressure, vascular tone, adipogenesis, along with glucose homeostasis and insulin signaling (9, 12–14). Based on this, we hypothesized that the oxylipin profiling could provide a unique mechanistic signature of MetS. A high-throughput validated mass spectrometry method allowing the quantitative profiling of over 130 oxylipins (15, 16) was applied to identify the oxylipin signature of MetS in a case-control study nested in the Polish branch of the Prospective Urban and Rural Epidemiological (PURE) cohort. We then replicated the study in an independent population, namely the French NutriNet-Santé cohort, allowing external validation of the newly discovered oxylipin signature.

## 2. Material and methods

### 2.1. Discovery and Replication cohorts

The discovery cohort was the Polish branch of the Prospective Urban and Rural Epidemiological cohort (n = 2036, aged 30-70 y) launched in 2009 (17). For the Polish branch of the PURE cohort, the recruitment of participants took place between 2007 and 2010. During the visit, socio-demographic (i.e. education, localization) and lifestyle questionnaires (including smoking habits, physical activity), medical tests (i.e. electrocardiogram, blood pressure), anthropometric measurements (i.e. weight, height, waist and hips circumference) and biochemical tests (fasting blood glucose, total cholesterol, HDL cholesterol (HDLc), LDL cholesterol (LDLc) and triglycerides (TG)) were realized. Moreover, at baseline, a 134 item food frequency questionnaire (FFQ) was completed by each participant. The study was approved by the Institutional Review Board of the Wroclaw Medical University (IRB number: KB-443/2006).

Independent external replication was conducted with participants from the web-based prospective French Nutrinet-Santé cohort (18) launched in 2014 and still ongoing (www.etude-nutrinet-sante.fr). The cohort included 160 000 participants (aged ≥18 y) but for the independent external validation the selection was made only from the participants having blood specimens (n =19 772, aged 30-70 y). Participants of the French Nutrinet-Santé cohort are asked to complete every year questionnaires to collect information regarding socio-demography (education and localization), lifestyle (including smoking habits, physical activity), health (disease history, menopausal status, anthropometric self-assessment) and dietary habits (3 x 24h dietary records including >3300 food items). Blood samples were collected at baseline and were used for the biochemical tests (fasting blood glucose, total cholesterol, HDLc, LDLc and TG). These selected participants also had a clinical examination including the assessment of blood pressure and anthropometric measurements (weight, height, waist and hip perimeters, fat mass, fat mass on body trunk and visceral fat). The study was approved by the International Research Board of the French Institute of Health and Medical research (IRB Inserm n°0000388FWA00005831) and the “Comité National Informatique et Liberté” (CNIL n°908450 and n°909216).

### 2.2. Study design and selection of participants

From the Discovery cohort (i.e. Polish branch of the PURE cohort) and the Replication cohort (i.e. French Nutrinet-Santé cohort), cross-sectional case-control studies were performed following similar designs. The outcome of the studies presented here was the metabolic syndrome (MetS) as defined by Alberti et al (3). According to this definition, participants were considered as a Case (i.e. diagnosed as having MetS) if they had at least three of the following criteria: elevated waist circumference (≥ 94 cm for men and 80 cm for women), elevated TG (≥ 150 mg/dL), reduced HDLc (< 40mg/dL for men and < 50 mg/dL for women), elevated blood pressure (systolic ≥ 130 and/or diastolic ≥ 85 mmHg) and elevated fasting glucose (≥ 100 mg/dL). Participants receiving pharmacotherapy for elevated TG, elevated blood pressure or hyperglycemia were considered as meeting the aforementioned criteria (3). Participants having less than three of the aforementioned criteria were considered as a Control. These studies are case-control studies aiming at comparing participants with diagnosed MetS (i.e. ≥3 criteria) vs Control participants (<3 criteria) (**Supplemental Figure S1**). To select the study samples, criteria of exclusion were similar between the Discovery and the Replication studies and included: participants diagnosed with cancer before and one year after blood draw, participants with cardiovascular events before blood draw (stroke, angina and heart failure), and participants with missing data for the cardiometabolic parameters (i.e. waist circumference, blood pressure, triglycerides, HDLc, fasting glucose). For the Discovery study, Case and Control participants were matched according to their sex, age (2 y classes), smoking status (never+former *vs* current) and physical activity (low *vs* moderate+intense). For the Replication study, matching factors included: sex, age (2 y classes), smoking status (never/former/irregular/current), physical activity (low/moderate/intense), menopausal status (not applicable/yes/no) and season of blood draw (winter/spring/summer/fall). To avoid the selection of incomparable Control participants between the two cohorts, the Control groups of each study were balanced according to the number of cardiometabolic criteria (i.e. 0, 1 or 2). At completion of the selection and matching processes, the Discovery study included 137 Cases and 137 Controls and the Replication cross-sectional study included 101 Cases and 101 Controls (**Supplemental Figure S1**).

### 2.3. Oxylipin and fatty acid quantification

For each participant, fasting blood was collected into EDTA and used to prepare plasma. Briefly, for the PURE participants, blood samples were directly centrifuged and plasma was either stored at -20°C for up to 3 days and then stored at -80°C or directly stored at -80°C. For the Nutrinet-Santé participants, blood samples were stored at +4°C for up to 24 hours and then centrifuged and plasma stored at -80°C. Of note, the conditions of transitory storage usually only affect a very limited number of oxylipins (19) and the samples used in these studies had never been thawed during their storage.

#### 2.3.1. Extraction and MS quantitative profiling of oxylipins

Samples collected in the Discovery and Replication cohorts were prepared and analyzed in two different laboratories but following a harmonized standard operating procedure known to generate very comparable results (15). All sample analysis was performed in a blinded fashion with regards to subject health status. Samples were randomized. The LC-MS method is described in detail in Ostermann et al. (20), Rund et al. (16) and Kutzer et al. (21). Quality controls (QC, i.e. intra-batch QC consisting of a plasma included in each batch of sample preparation and inter-batch QC consisting of a pool of oxylipin extracts) were prepared to control for potential bias due to sample preparation or analytical drifts. Briefly, 10 µL of an antioxidant mixture including butylated hydroxytoluene (BHT, 0.2 mg/mL), *trans*-AUCB (100 µM), indomethacin (100 µM) and 10 µL of an internal standard mixture (including 20 deuterated standards: 100 nM of each ^2^H_4_-8-iso-PGF2α, ^2^H_4_-6-keto-PGF1α, ^2^H_4_-PGF2α, ^2^H_11_-8,12-iso-iPF2α-VI, ^2^H_4_-PGB2, ^2^H_5_-LxA4, ^2^H_5_-RvD1, ^2^H_5_-RvD2, ^2^H_4_-LTB4, ^2^H_4_-9,10-DiHOME, ^2^H_11_-11,12-DiHETrE, ^2^H_4_-13-HODE, ^2^H_4_-9-HODE, ^2^H_6_-20-HETE, ^2^H_8_-15-HETE, ^2^H_8_-12-HETE, ^2^H_8_-5-HETE, ^2^H_4_-12(13)-EpOME, ^2^H_11_-14(15)-EpETrE, ^2^H_11_-8(9)-EpETrE in MeOH) were added to 100 µL of plasma. Following protein precipitation with 400 µL of cold isopropanol (LC-MS grade, Fisher Scientific) and the hydrolysis of esterified oxylipins with 0.6 M of potassium hydroxide (MeOH/H_2_O, 75/25, *v/v*, 60°C, 30 min), total oxylipins (i.e. free and esterified) were extracted using Bond Elut Certify II SPE cartridges (200 mg, 3 mL, Agilent Technologies). Oxylipins were eluted into glass tubes containing 10 µL of 30% glycerol in MeOH using ethyl acetate/*n*-hexane/acetic acid (75/25/1, *v/v/v*). Samples were evaporated and the residue was reconstituted in 50 µL MeOH and stored at -80°C before mass spectrometry (MS) analysis. Extracted oxylipins were measured using electrospray ionization in negative ion mode and multiple reaction monitoring (MRM) using the most abundant and specific precursor ion/product ion transitions to build an acquisition method capable of detecting 133 analytes and 20 isotope-labelled internal standards. The ion spray voltage was set at −4500 V at a temperature of 40°C. Collisional activation of the oxylipin precursor ions was achieved with nitrogen as the collision gas with the declustering potential, entrance potential, and collision energy optimized for each metabolite (16, 19–21). Oxylipins were identified by matching their MRM signal and chromatographic retention time with those of pure standards. Absolute concentrations are reported in the **Supplemental Table S1**.

#### 2.3.2. Fatty acid profiling

Plasma samples collected in the Discovery and Replication cohorts were prepared and analyzed in two different laboratories as described in Lillington et al. (22). Briefly, 10 µL of an internal standard (TG19) were added to 10 µL of plasma. Following the hydrolysis with 1 mL KOH (0.5 M), the derivatization with 1 mL heptane and 1 mL BF_3_-MeOH (80°C, 1h) and the addition of 1 mL H_2_O, the fatty acids methyl esters (FAMEs) were extracted with 1 mL H_2_O and 2 mL of heptane, dried and dissolved in 20 µL of ethyl acetate. FAMEs were separated by gas chromatography (GC) on a Clarus 600 Perkin Elmer system using a Famewax RESTEK fused silica capillary column (30 m x 0.32 mm, 0.25 µm film thickness). Oven temperature was programmed from 110°C to 220°C at a rate of 2°C/min and the carrier gas was hydrogen (0.5 bar). The injector and detector temperatures were at 225°C and 245°C, respectively. All of the quantitative calculations were based on the chromatographic peak area relative to the internal standard. Using these methods, the number of detected and accurately quantified fatty acids was as follows for the different studies: 21 fatty acids for the Discovery study (i.e. PURE) and 19 fatty acids oxylipins for the Replication cross-sectional study (i.e. Nutrinet-Santé). Absolute concentrations are reported in **Supplemental Table S1**.

### 2.4. Statistical analysis

#### 2.4.1. Metadata statistical analysis

Metadata associated with each participant selected in the Discovery (i.e. PURE) and the Replication (i.e. Nutrinet-Santé) studies included qualitative data (i.e. sex, smoking, education, localization, season of blood draw) and quantitative data (i.e. weight, body mass index (BMI), waist and hip circumference, systolic and diastolic blood pressure, fasting blood glucose, total cholesterol, TG, HDLc and LDLc, alternative healthy eating index (AHEI) score and plasma fatty acids) recorded or assessed at baseline. The AHEI score was calculated to estimate the quality of diet of participants and based on ten components reflecting recent dietary guidelines (23). Based on the cardiometabolic criteria, the metabolic score (MetS-z-score) was calculated as described in https://github.com/metscalc/metscalc/ as follows: MetS-z-score = Y + a*waist - b*HDLc + c*SBP + d*log(TG) + e*glucose, with the coefficients Y, a, b, c, d and e being determined according sex, ethnicity and age (adults vs teenager). This derived z-score is a continuous variable providing an integrative assessment of the cardiometabolic status (24). Differences between Cases and Controls for each parameter were assessed using univariate analysis and taking into consideration the matching of participants. The non-parametric Wilcoxon signed-rank test was used for quantitative variables whereas the contingency Fisher’s Exact test was used for qualitative variables. P-values were corrected for multiple testing using the false discovery rate correction of Benjamini-Hochberg (BH). Moreover, a special emphasis was put on the cardiometabolic parameters (i.e. waist perimeter, systolic and diastolic blood pressure, fasting glucose, TG, HDLc and MetS-z-score) through Volcano plot representations. Cardiometabolic variables were mean-centered and reduced allowing comparison as the difference in variables expressed in standard deviation between Case and Control participants. Each variable was represented in the Volcano plot with the difference in standard deviation (SD) between Case and Control participants in the x-axis and the -log(p-value) of the Wilcoxon signed-rank test (BH-corrected) in the y-axis. Statistical analysis and graphical representations were generated by the R statistical computing environment (https://www.R-project.org/) using the “EnhancedVolcano” package (https://github.com/kevinblighe/EnhancedVolcano).

#### 2.4.2. Pre-processing of MS oxylipin data

MS data generated from the Discovery and the Validation studies were pre-processed following the same protocol as described in the Supplemental method document. This includes detailed information regarding MS data integration, normalization and imputation as well as information regarding data adjustment. This latter pre-processing step aims to reduce the impact of total oxylipin levels on data variability and required discrete intensity adjustments as described in the Supplemental method. For the different studies, the number of quantified oxylipins was as follows: 88 oxylipins for the Discovery study (i.e. PURE) and 58 oxylipins for the Replication study (i.e. Nutrinet-Santé). Merging the oxylipin data matrix generated in the Discovery and the Replication studies identified 54 oxylipins in common. This common matrix of 54 oxylipins was used for the selection and validation of oxylipins thereafter highlighted as ‘candidates’ (shared upon request by the corresponding author).

#### 2.4.3. Candidate oxylipins selection

To leverage the case-control design of our studies and the matching of the selected participants, conditional logistic regression was used to model the association of a set of oxylipins with the odds of having a MetS diagnosis. Moreover, to overcome the high-dimensional setting of our datasets and to put oxylipins unlinked to the outcome modeled aside, the Elastic-net penalization (25) method was applied for the initial oxylipin selection. This method relies on a penalization parameter that controls for the strength of selection by shrinking the coefficients toward zero (setting some to exactly zero thus performing variable selection). Contrary to methods as the least absolute shrinkage and selection operator (LASSO) penalization that tend to select only one representative from a pool of multi-collinear variables, Elastic-net penalization preserves all variables appearing linked to the outcome. This was considered to be important for the biological interpretation since even if oxylipins are collinear they may bring complementary information in the interpretation process. These penalization models were computed using the R software with OPT2D function from the “penalized” package (26). As penalized regression may lead to unstable solutions due to its cross-validation-based parameter determination process, bootstrap resampling was used to enhance the robustness of oxylipin selection. Elastic-net-penalized conditional logistic regressions were repeated on 350 bootstrapped samples; the oxylipins were ordered by decreasing percentage of selection across bootstraps and the oxylipin signature of metabolic syndrome was focused on the oxylipins selected in ≥80% of bootstraps (“candidate oxylipins”) for the Discovery and Replication studies. A scoring of analytical robustness was established for each candidate oxylipin selected taking into account previously published results regarding (i) the stability during transitory and long term storage (19) and (ii) the technical and interlaboratory variabilities (15). The scoring also takes into consideration the percentage of missing data imputation. Oxylipins with putative low score of analytical robustness were set apart from the initial selection. The process of candidate oxylipin selection was independently duplicated in the Discovery and Replication studies in order to (i) assess the consistency of the selected oxylipins and (ii) generate a complete list of oxylipins including common and population-specific oxylipins.

#### 2.4.4. Model construction and validation

The objective was to evaluate whether a discrimination model fitted on the Discovery study could be efficient on an independent study regarding the outcome of prediction. First and foremost, in order to merge the oxylipin dataset generated in the Discovery and the Replication studies, a harmonization of the quantitative data adjustment protocol was necessary (see supplemental methods). Then, a LASSO-penalized conditional logistic regression model was constructed using the participants of the Discovery study and including the oxylipins previously selected in the Discovery and in the Replication studies. The LASSO penalization was chosen to remove putative redundant oxylipins from the complete signature, with model classification performances assessed for both Discovery and Replication study participants using the Area Under the Receiver Operating Characteristic Curves (AUC_ROC_) which were computed using 10-fold cross-validation, with confidence intervals and cross-validated error rates being calculated. Analysis and representation were performed by the R software using the “glmnet” (27), “pROC” (28) and “selectiveInference” (29) packages. Odds ratios of each oxylipin of the optimized signature were estimated and plotted as Circos plot (30).

#### 2.4.5. OxyScore calculation

Based on the LASSO-penalized conditional logistic regression models created in the Discovery study and validated in the Replication study, the probability of being a Case (coined OxyScore) was calculated for each participant as follows: 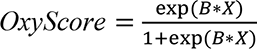 where (B*X) corresponds to a combination of b _0_*x_0_, b_1_*x_1_, …, b_p_*x_p_ (p the number of oxylipins in the model) with b_n_ being the adjusted coefficient of the n-th oxylipin in the LASSO regression model and x_n_ the corresponding oxylipin concentration.

#### 2.4.6. OxyScore correlation

In order to assess the consistency and the complementarity of the OxyScore with the clinical criteria of metabolic syndrome, Spearman correlations were established between the OxyScore calculated for each participant of the Discovery study and the criteria of MetS (including the MetS-z-score). Correlations were considered as significant when p <0.05 and r ≥0.5 (noteworthy link) or r ≥0.7 (strong link).

#### 2.4.7. OxyScore adjustment

In order to see the impact of cardiometabolic criteria in the performances of the OxyScore and in the contribution of each oxylipin to the OxyScore, the LASSO-penalized conditional logistic regression model created in the Discovery study and validated in the Replication study (used for the calculation of the OxyScore) was adjusted with each cardiometabolic criterion (i.e. waist circumference, systolic and diastolic blood pressure, fasting glucose, TG, HDLc and BMI) one by one. Performance of the adjusted OxyScore was assessed as previously described (i.e. AUC_ROC_ computed using the 10-fold cross-validation, confidence intervals and cross-validated error rate). Odds ratios of each oxylipin of the adjusted OxyScore were estimated. The impact of adding the criteria of MetS into the LASSO model on the maintenance or exclusion of the oxylipins of the model was evaluated.

#### 2.4.8. Univariate analysis

To determine which oxylipins of the initial signature could reflect the outcome individually, Wilcoxon signed-rank test with BH multiple-tests correction were computed for the Discovery and Replication cross-sectional studies.

#### 2.4.9. Study approval

Participants provided written consent to be included in the Polish branch of the PURE cohort or the web-based prospective French Nutrinet-Santé cohort. These were approved by the Institutional Review Board of the Wroclaw Medical University (IRB number: KB-443/2006) and by the International Research Board of the French Institute of Health and Medical research (IRB Inserm n°0000388FWA00005831) and the “Comité National Informatique et Liberté” (CNIL n°908450 and n°909216).

## 3. Results

### 3.1. Baseline characteristics of participants

As shown in **Table 1**, differences between Cases (i.e. participants with ≥3 criteria of MetS including obesity, high blood pressure, hypertriglyceridemia, low HDL-c and hyperglycemia as describe in (3)) and Controls (participants with <3 criteria of MetS) were comparable for both the Discovery and the Replication studies, except for the level of education, the localization and the level of low density lipoprotein cholesterol (LDLc) that were significantly different only between Case and Control participants of the Discovery study. Moreover, when looking at deviation from the mean of the cardiometabolic criteria (**Figure 1**), differences between Case and Control participants were slightly more pronounced in the Discovery study than in the Replication study. The contrast between both studies was particularly apparent for the MetS-z-score (z-score integrating the five criteria of MetS (24)) and HDLc. When comparing Case and Control participants across studies, there were several significant differences, especially between the Case participants (last column, **Table 1**). Notably, in comparison with the Discovery study, participants of the Replication study were significantly older (61 y *vs* 54 y, on average, both in cases and controls). Significant differences between studies were also observed for all lifestyle variables measured. Concerning the clinical variables of the Case participants, they were significantly higher in the Discovery study with the exception of cholesterol (total, HDLc and LDLc). Regarding the MetS components, their relative distributions for the Control and Case participants were very similar across studies except for the proportion of Control participants with high blood pressure that was significantly higher in the Discovery study in comparison with the Control participants of the Replication study (i.e. 65.0% vs 49.5%). The Discovery study also included more Case participants with hyperglycemia (75.2%) than the Replication study (62.4%).

**Table 1.**
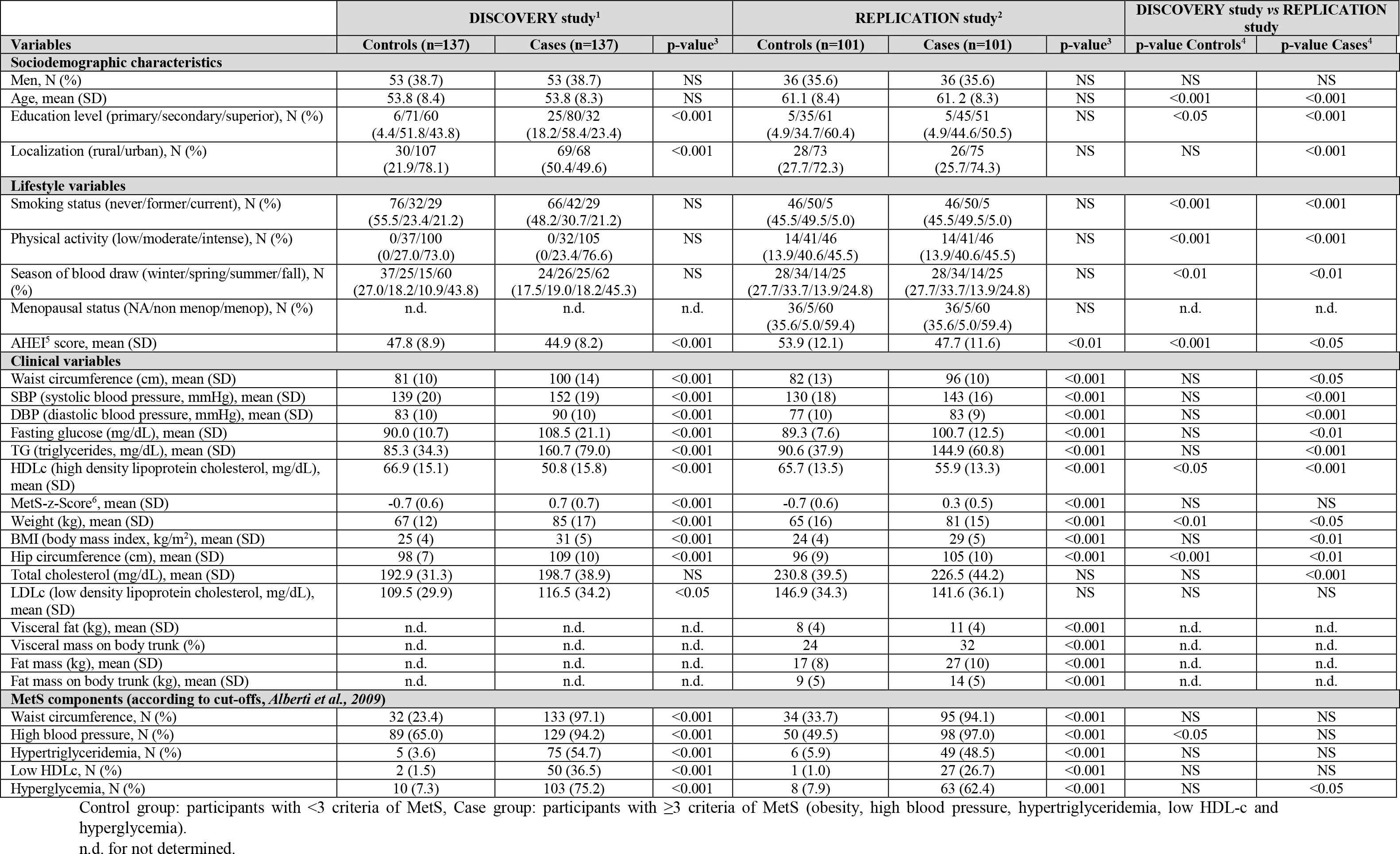
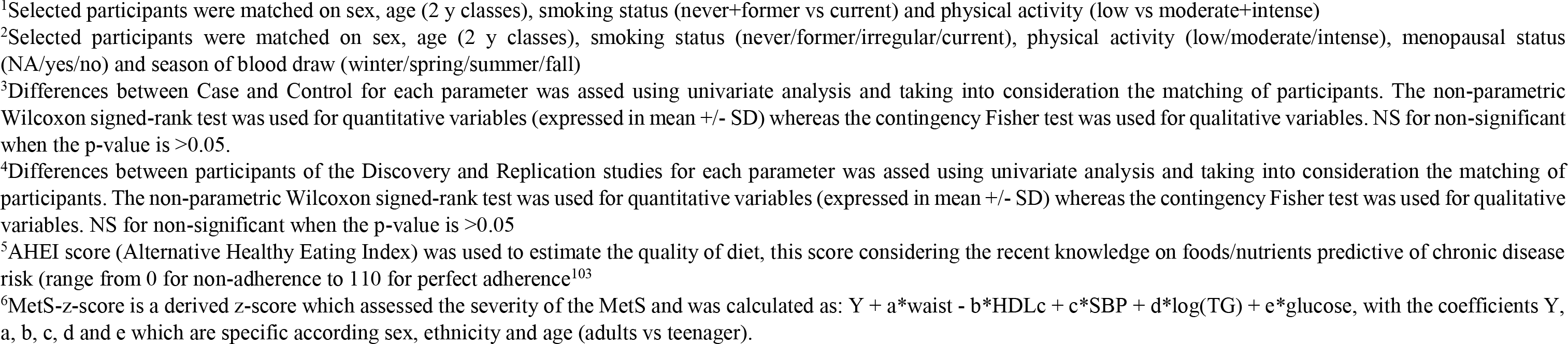
Baseline characteristics of the population selected for the Discovery and Replication studies.

|  | DISCOVERY study<sup>1</sup> |  |  | REPLICATION study<sup>2</sup> |  |  | DISCOVERY study vs REPLICATION study |  |
| --- | --- | --- | --- | --- | --- | --- | --- | --- |
| Variables | Controls (n=137) | Cases (n=137) | p-value<sup>3</sup> | Controls (n=101) | Cases (n=101) | p-value<sup>3</sup> | p-value Controls<sup>4</sup> | p-value Cases<sup>4</sup> |
| **Sociodemographic characteristics** |  |  |  |  |  |  |  |  |
| Men, N (%) | 53 (38.7) | 53 (38.7) | NS | 36 (35.6) | 36 (35.6) | NS | NS | NS |
| Age, mean (SD) | 53.8 (8.4) | 53.8 (8.3) | NS | 61.1 (8.4) | 61.2 (8.3) | NS | <0.001 | <0.001 |
| Education level (primary/secondary/superior), N (%) | 6/71/60<br>(4.4/51.8/43.8) | 25/80/32<br>(18.2/58.4/23.4) | <0.001 | 5/35/61<br>(4.9/34.7/60.4) | 5/45/51<br>(4.9/44.6/50.5) | NS | <0.05 | <0.001 |
| Localization (rural/urban), N (%) | 30/107<br>(21.9/78.1) | 69/68<br>(50.4/49.6) | <0.001 | 28/73<br>(27.7/72.3) | 26/75<br>(25.7/74.3) | NS | NS | <0.001 |
| **Lifestyle variables** |  |  |  |  |  |  |  |  |
| Smoking status (never/former/current), N (%) | 76/32/29<br>(55.5/23.4/21.2) | 66/42/29<br>(48.2/30.7/21.2) | NS | 46/50/5<br>(45.5/49.5/5.0) | 46/50/5<br>(45.5/49.5/5.0) | NS | <0.001 | <0.001 |
| Physical activity (low/moderate/intense), N (%) | 0/37/100<br>(0/27.0/73.0) | 0/32/105<br>(0/23.4/76.6) | NS | 14/41/46<br>(13.9/40.6/45.5) | 14/41/46<br>(13.9/40.6/45.5) | NS | <0.001 | <0.001 |
| Season of blood draw (winter/spring/summer/fall), N (%) | 37/25/15/60<br>(27.0/18.2/10.9/43.8) | 24/26/25/62<br>(17.5/19.0/18.2/45.3) | NS | 28/34/14/25<br>(27.7/33.7/13.9/24.8) | 28/34/14/25<br>(27.7/33.7/13.9/24.8) | NS | <0.01 | <0.01 |
| Menopausal status (NA/non menop/menop), N (%) | n.d. | n.d. | n.d. | 36/5/60<br>(35.6/5.0/59.4) | 36/5/60<br>(35.6/5.0/59.4) | NS | n.d. | n.d. |
| AHEI<sup>5</sup> score, mean (SD) | 47.8 (8.9) | 44.9 (8.2) | <0.001 | 53.9 (12.1) | 47.7 (11.6) | <0.01 | <0.001 | <0.05 |
| **Clinical variables** |  |  |  |  |  |  |  |  |
| Waist circumference (cm), mean (SD) | 81 (10) | 100 (14) | <0.001 | 82 (13) | 96 (10) | <0.001 | NS | <0.05 |
| SBP (systolic blood pressure, mmHg), mean (SD) | 139 (20) | 152 (19) | <0.001 | 130 (18) | 143 (16) | <0.001 | NS | <0.001 |
| DBP (diastolic blood pressure, mmHg), mean (SD) | 83 (10) | 90 (10) | <0.001 | 77 (10) | 83 (9) | <0.001 | NS | <0.001 |
| Fasting glucose (mg/dL), mean (SD) | 90.0 (10.7) | 108.5 (21.1) | <0.001 | 89.3 (7.6) | 100.7 (12.5) | <0.001 | NS | <0.01 |
| TG (triglycerides, mg/dL), mean (SD) | 85.3 (34.3) | 160.7 (79.0) | <0.001 | 90.6 (37.9) | 144.9 (60.8) | <0.001 | NS | <0.001 |
| HDLc (high density lipoprotein cholesterol, mg/dL), mean (SD) | 66.9 (15.1) | 50.8 (15.8) | <0.001 | 65.7 (13.5) | 55.9 (13.3) | <0.001 | <0.05 | <0.001 |
| MetS-z-Score<sup>6</sup>, mean (SD) | -0.7 (0.6) | 0.7 (0.7) | <0.001 | -0.7 (0.6) | 0.3 (0.5) | <0.001 | NS | NS |
| Weight (kg), mean (SD) | 67 (12) | 85 (17) | <0.001 | 65 (16) | 81 (15) | <0.001 | <0.01 | <0.05 |
| BMI (body mass index, kg/m<sup>2</sup>), mean (SD) | 25 (4) | 31 (5) | <0.001 | 24 (4) | 29 (5) | <0.001 | NS | <0.01 |
| Hip circumference (cm), mean (SD) | 98 (7) | 109 (10) | <0.001 | 96 (9) | 105 (10) | <0.001 | <0.001 | <0.01 |
| Total cholesterol (mg/dL), mean (SD) | 192.9 (31.3) | 198.7 (38.9) | NS | 230.8 (39.5) | 226.5 (44.2) | NS | NS | <0.001 |
| LDLc (low density lipoprotein cholesterol, mg/dL), mean (SD) | 109.5 (29.9) | 116.5 (34.2) | <0.05 | 146.9 (34.3) | 141.6 (36.1) | NS | NS | NS |
| Visceral fat (kg), mean (SD) | n.d. | n.d. | n.d. | 8 (4) | 11 (4) | <0.001 | n.d. | n.d. |
| Visceral mass on body trunk (%) | n.d. | n.d. | n.d. | 24 | 32 | <0.001 | n.d. | n.d. |
| Fat mass (kg), mean (SD) | n.d. | n.d. | n.d. | 17 (8) | 27 (10) | <0.001 | n.d. | n.d. |
| Fat mass on body trunk (kg), mean (SD) | n.d. | n.d. | n.d. | 9 (5) | 14 (5) | <0.001 | n.d. | n.d. |
| **MetS components (according to cut-offs, Alberti et al., 2009)** |  |  |  |  |  |  |  |  |
| Waist circumference, N (%) | 32 (23.4) | 133 (97.1) | <0.001 | 34 (33.7) | 95 (94.1) | <0.001 | NS | NS |
| High blood pressure, N (%) | 89 (65.0) | 129 (94.2) | <0.001 | 50 (49.5) | 98 (97.0) | <0.001 | <0.05 | NS |
| Hypertriglyceridemia, N (%) | 5 (3.6) | 75 (54.7) | <0.001 | 6 (5.9) | 49 (48.5) | <0.001 | NS | NS |
| Low HDLc, N (%) | 2 (1.5) | 50 (36.5) | <0.001 | 1 (1.0) | 27 (26.7) | <0.001 | NS | NS |
| Hyperglycemia, N (%) | 10 (7.3) | 103 (75.2) | <0.001 | 8 (7.9) | 63 (62.4) | <0.001 | NS | <0.05 |

Control group: participants with <3 criteria of MetS, Case group: participants with ≥3 criteria of MetS (obesity, high blood pressure, hypertriglyceridemia, low HDL-c and hyperglycemia).
n.d. for not determined.
<sup>1</sup>Selected participants were matched on sex, age (2 y classes), smoking status (never+former vs current) and physical activity (low vs moderate+intense)
<sup>2</sup>Selected participants were matched on sex, age (2 y classes), smoking status (never/former/irregular/current), physical activity (low/moderate/intense), menopausal status (NA/yes/no) and season of blood draw (winter/spring/summer/fall)
<sup>3</sup>Differences between Case and Control for each parameter was assessed using univariate analysis and taking into consideration the matching of participants. The non-parametric Wilcoxon signed-rank test was used for quantitative variables (expressed in mean +/- SD) whereas the contingency Fisher test was used for qualitative variables. NS for non-significant when the p-value is >0.05.
<sup>4</sup>Differences between participants of the Discovery and Replication studies for each parameter was assessed using univariate analysis and taking into consideration the matching of participants. The non-parametric Wilcoxon signed-rank test was used for quantitative variables (expressed in mean +/- SD) whereas the contingency Fisher test was used for qualitative variables. NS for non-significant when the p-value is >0.05
<sup>5</sup>AHEI score (Alternative Healthy Eating Index) was used to estimate the quality of diet, this score considering the recent knowledge on foods/nutrients predictive of chronic disease risk (range from 0 for non-adherence to 110 for perfect adherence)<sup>103</sup>
<sup>6</sup>MetS-z-score is a derived z-score which assessed the severity of the MetS and was calculated as: $Y + a \cdot \text{waist} - b \cdot \text{HDLc} + c \cdot \text{SBP} + d \cdot \log(\text{TG}) + e \cdot \text{glucose}$ , with the coefficients Y, a, b, c, d and e which are specific according sex, ethnicity and age (adults vs teenager).

**Figure 1.**
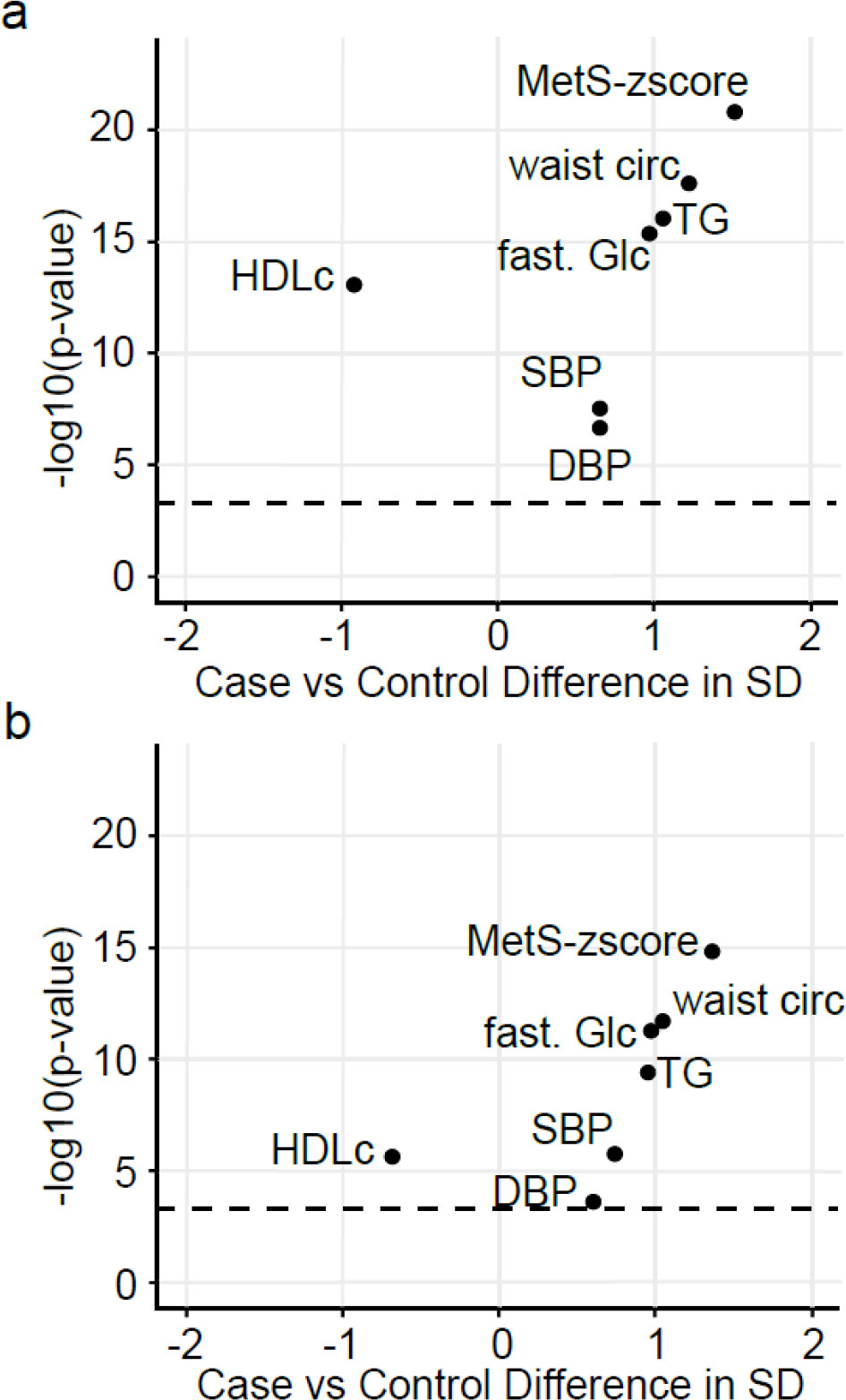
Baseline cardiometabolic differences between the Control and MetS participants of the Discovery and the Replication studies. Volcano plots showing baseline differences of MetS status (**a**) between Case and Control participants of the Discovery study and (**b**) between Case and Control participants of the Replication study. Differences between Case and Control are expressed in standard deviation (SD) and were assessed using Wilcoxon signed-rank test followed by multiple tests correction (BH). Dash line represents the significant threshold (p-value BH =0·0005). HDLc: high density lipoprotein cholesterol, TG: triglycerides, fast. Glc: fasting glucose, waist circ: waist circumference, DBP and SBP: diastolic and systolic blood pressure.

### 3.2. Lipidomic profiling identifies consistent oxylipin signatures of metabolic syndrome between the Discovery and the Replication studies

In order to validate the consistency of the oxylipin signature discriminating participants with or without MetS, we performed two independent oxylipin selections in the Discovery and Replication studies respectively (**Supplemental Figure S2**). The bootstrap-enhanced Elastic-net selection made in the Discovery study identified 28 candidate oxylipins (i.e. oxylipins found in ≥80% of bootstraps; **Supplemental Figure S2a**). A scoring of analytical robustness was established for each candidate oxylipin selected taking into account previously published results regarding (i) the stability during transitory and long term storage (19) and (ii) the technical and interlaboratory variabilities (15). The scoring also takes into consideration the percentage of missing data imputation. Using this scoring system, 5 out of the 28 candidate oxylipins were excluded from further steps (namely 12,13-DiHOME, 15(16)-EpODE, 9-HETE, 14(15)-EpETrE, and 19,20-DiHDPE) because of putative low analytical robustness. The 23 remaining candidate oxylipins selected in the Discovery study include 4 epoxy-PUFAs (i.e. 9(10)-epoxy-stearic acid, 9(10)-EpOME, 11(12)-EpETrE and 11(12)-EpETE), 2 ketone-PUFAs (9- and 13-oxo-ODE), 12 mid-chain hydroxyl-PUFAs (10- and 12-HODE, 5- and 15-HETrE, 5-, 15- and 16-HETE, 5- and 8-HEPE, 4-, 7- and 11-HDHA) and 5 vicinal dihydroxy-PUFAs (9,10-DiHOME, 9,10-DiHODE, 5,6- and 14,15-DiHETrE, and 7,8-DiHDPE). The replication of the bootstrap-enhanced Elastic-net selection in the Replication study identified 19 candidate oxylipins (**Supplemental Figure S2b**), two of them (i.e. 9(10)- and 15(16)-EpODE) being put aside because of putative low analytical robustness using the same scoring system mentioned above. The 17 remaining candidate oxylipins selected in the Replication study include 5 epoxy-PUFAs (9(10)-epoxy-stearic acid, 12(13)-EpODE, 14(15)-EpEDE, 11(12)-EpETrE and 11(12)-EpETE), 2 ketone- PUFAs (9- and 13-oxo-ODE), 8 mid-chain hydroxy-PUFAs (9-, 13- and 15-HODE, 5- and 12-HETrE, 5- HEPE, 7- and 11-HDHA) and 2 vicinal dihydroxy-PUFAs (9,10-DiHODE and 5,6-DiHETrE). Among these 17 oxylipins, 11 were in common with the 23 candidate oxylipins already selected in the Discovery study (**Figure 2**) supporting the consistency of the identified oxylipin signature. Thus 29 oxylipins were identified as potential biomarkers of MetS from the two studies.

**Figure 2.**
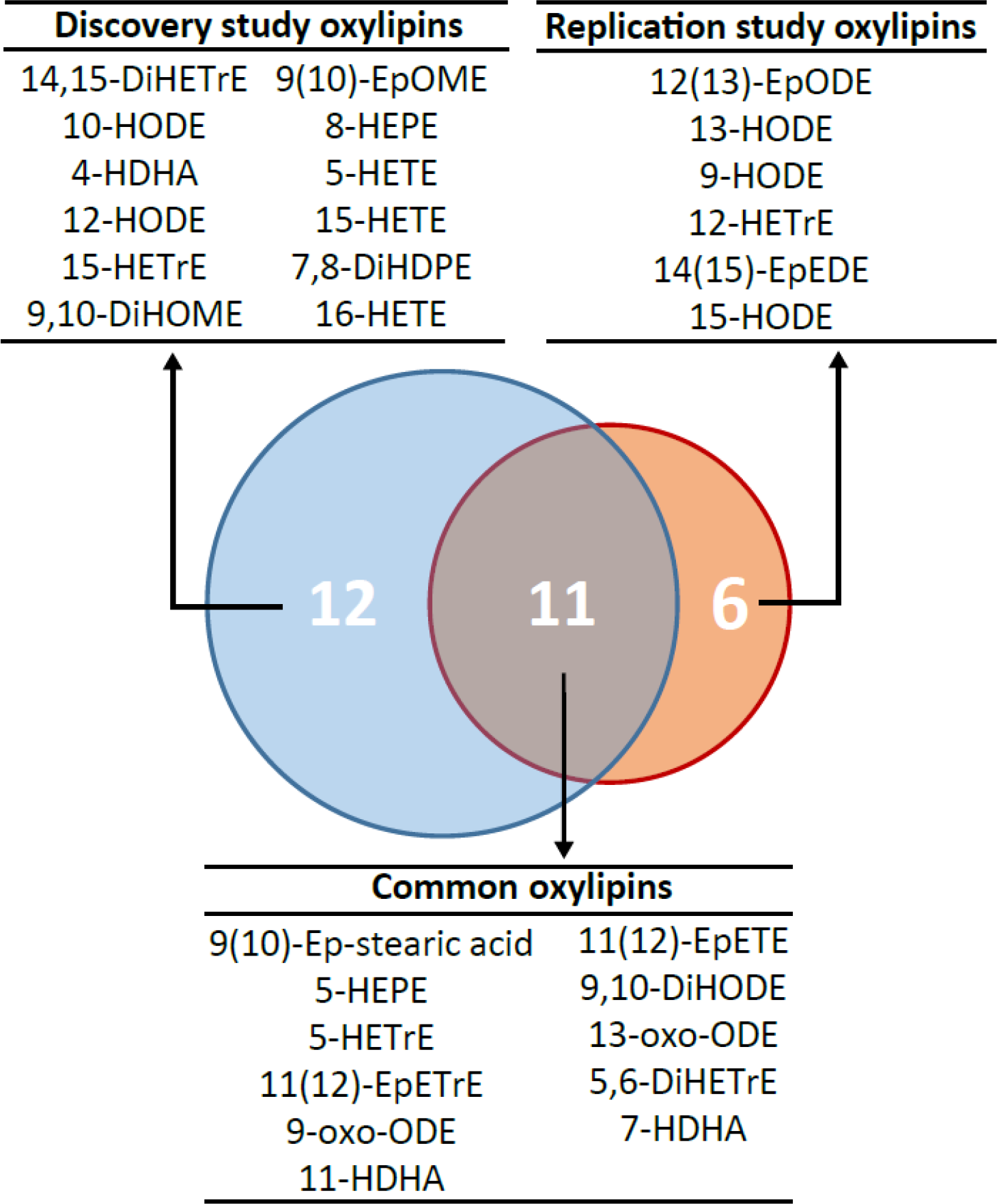
Consistency of the oxylipin signature discriminating MetS participants independently selected in the Discovery and the Replication studies. Venn diagram showing the common and specific oxylipins selected in the Discovery and Replication studies.

### 3.3. The identified candidate oxylipins provide a unique mechanistic phenotype of cardiometabolic health

The 29 identified candidate oxylipins (highlighted in red, green or grey, **Figure 3**) include 12 octadecanoids, 13 eicosanoids and 4 docosanoids (i.e. oxylipins respectively derived form 18-, 20- or 22-carbons PUFAs). Among them, 18 were derived from omega-6 PUFAs (including 9 from linoleic acid (LA/C18:2ω6), 3 from dihomo-γ-linolenic acid (DGLA/C20:3ω6) and 6 from arachidonic acid (AA/C20:4ω6), **Figure 3a**), 9 from omega-3 PUFAs (including 2 from α-linolenic acid (ALA/C18:3ω3), 3 from eicosapentaenoic acid (EPA/C20:5ω3) and 4 from docosahexaenoic acid (DHA/C22:6ω3), **Figure 3b**), and 2 from oleic acid (OA, C18:1ω9, **Figure 3c**). Regarding the pathways of biosynthesis, 3 candidate oxylipins (8-HEPE, 10- and 12- HODE) arise from the free-radical mediated pathway, 13 (9- and 13-HODE and their oxo-derivatives, 5-, 12- and 15-HETrE, 5- and 15-HETE, 5-HEPE, 4-, 7- and 11-HDHA) from enzymatic pathways involving the lipoxygenases (5-, 12 or 15-LOX) and 13 (9(10)-EpOME, 9,10-DiHOME, 15-HODE, 14(15)-EpEDE, 5,6- and 14,15-DiHETrE, 11(12)-EpETrE, 16-HETE, 9,10-DiHODE, 12(13)-EpODE, 11(12)-EpETE, 7,8-DiHDPE and 9(10)-Ep stearic acid) from the CYP pathway (cytochrome P450 and soluble epoxide hydrolase, sEH). Of note, some oxylipins may have dual origins: all LOX products (especially 9-HODE and 13-HODE) could also be produced by free-radical mediated reactions (31) while 12-HETrE could arise from 12-LOX or CYP (32). No prostanoids (i.e. cyclooxygenase-derived (COX-derived) eicosanoids) were included in the candidate oxylipins but it should be noted that alkaline conditions applied to liberate esterified oxylipins destroy the ω-hydroxy-keto prostanoids (e.g. PGEs, PGDs) and thromboxanes (20). When looking at the differences in concentration between cases and controls in the Discovery study (respectively highlighted in red or green for higher and lower concentration in Cases *vs* Controls, **Figure 3** **and Supplemental Table S1**), the first striking observation is related to the HODEs derived from the LOX and free-radical mediated pathways that were all significantly lower in those with MetS than in Controls. More precisely, 9-HODE, 13-HODE and their oxo-derivatives (9-oxo-ODE and 13-oxo-ODE) were lower by 11.0%, 22.2%, 49.4% and 34.1% respectively while the 10-HODE and 12-HODE (derived from singlet oxygen oxidation (33)) were lower by 21.4% and 42.7% respectively. In contrast, CYP-derived epoxy-PUFAs were systematically higher in the MetS participants in comparison with Controls, notably 14(15)- EpEDE, 12(13)-EpODE and 9(10)-epoxy-stearic acid that were significantly higher by 16.8%, 30.6% and 16.7% respectively. The concentrations of vicinal dihydroxy-PUFAs were mostly lower in MetS participants in comparison with Controls with 9,10-DiHOME, 14,15-DiHETrE and 7,8-DiHDPE being respectively lower by 15.7% (p<0.01), 5.1% (ns) and 30.4% (p<0.01). The 5-LOX pathway showed mixed and modest findings with two oxylipins tending to be higher (i.e. 5-HETrE and 7-HDHA) and three tending to be lower (i.e. 5-HETE, 5-HEPE and 4-HDHA) in MetS participants. The 12-LOX derivative of DGLA (i.e. 12-HETrE) also tended to be lower in MetS participants while 15-LOX derivatives of long chain PUFAs (i.e. 15-HETrE and 15-HETE) tended to be higher. In the Replication study, differences between those with MetS and Controls were similar for 19 oxylipins out of the 29 (i.e. 66%) to the differences observed in the Discovery study (**Supplemental Table S1**). Notably, except for 9(10)-EpOME, all epoxy-PUFAs were higher in MetS participants. However, results were different between the Discovery and Replication studies regarding ten candidate oxylipins (i.e. 34%). Among these, three LA metabolites (i.e. 9-HODE, 13-HODE, 9-oxo-ODE) changed in the opposite direction while 9(10)-EpOME tended to be higher in the MetS group. For the vicinal dihydroxy-PUFAs, the striking difference concerns 9,10-DiHODE that was lower by 41% (p<0.01) in MetS cases in comparison with Controls. For the 5-LOX pathway, there were higher concentrations of 2 oxylipins in MetS participants, namely 5-HETE (+9.8%, p<0.05) and 5-HEPE (+15.1%, p<0.05) and the difference in 5-HETrE became significant (+35.3%, p<0.001). Finally, the 12-LOX derivative of DGLA (i.e. 12-HETrE) and the CYP-ω metabolites (i.e. 15-HODE and 16-HETE) were found in lower (−31.8%, p<0.05) or higher concentration (+16.0%, p<0.05) in MetS participants.

**Figure 3.**
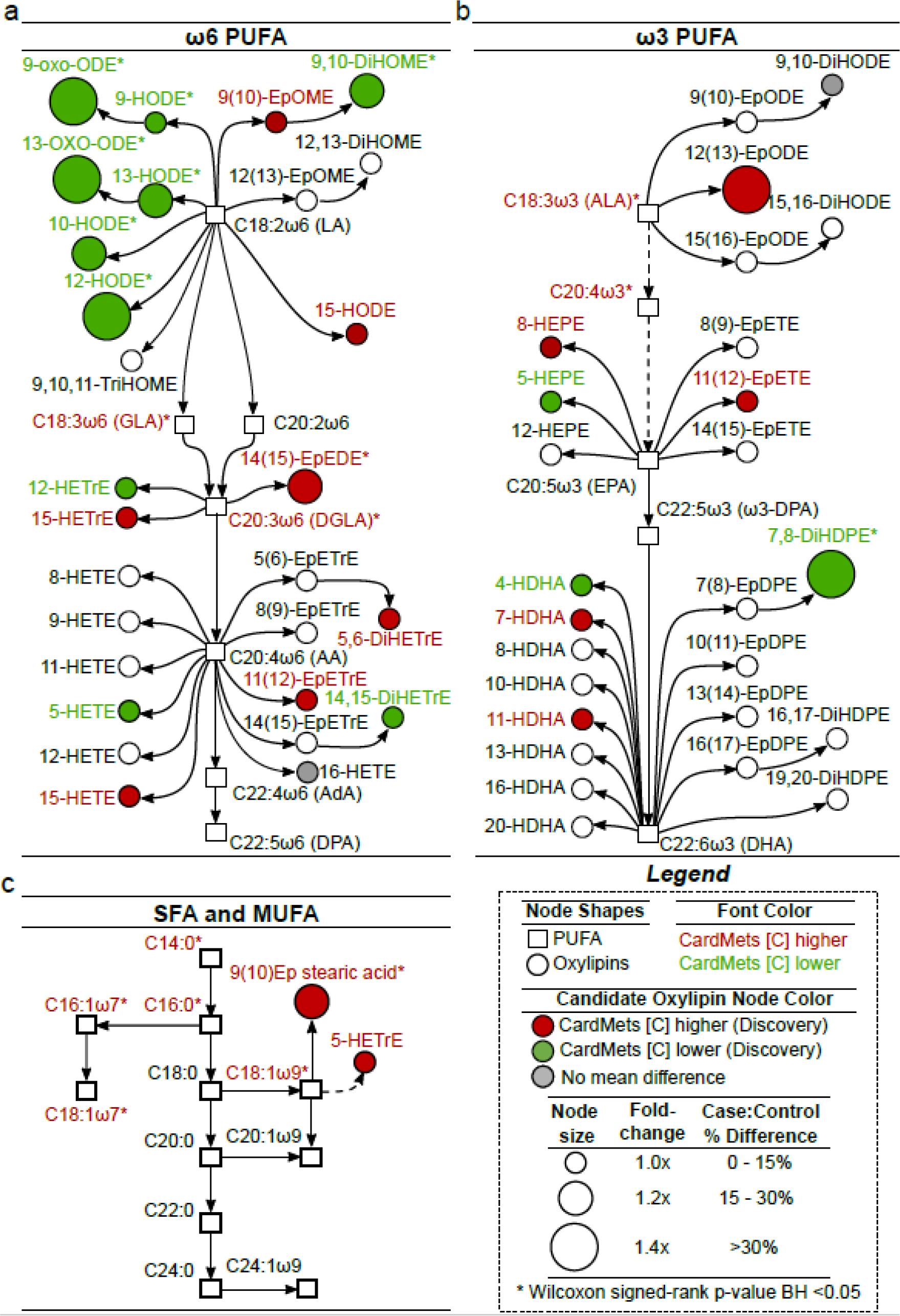
Biological origin of the candidate oxylipins discriminating participants with MetS. Metabolic pathway showing all oxylipins (n=54) and the fatty acids (n=25) (represented by circles and squares, respectively) measured in the Discovery and Replication cross-sectional studies. Pathways of oxylipins biosynthesis derived from omega-6 PUFAs (**a**), omega-3 PUFAs (**b**) and SFA/MUFAs (**c**). The oxylipins selected in the Discovery and Replication studies (candidates oxylipins n=29) are highlighted in color. Differences in concentration between controls and MetS participants in the Discovery study are highlighted in red, green or grey. Size of nodes represents the fold change between the two groups. Significant oxylipins from Wilcoxon signed-rank are represented by a star. LA: Linoleic Acid; GLA: γ-Linolenic Acid; DGLA: Dihomo-γ-Linolenic Acid; AA: Arachidonic Acid; AdA: Adrenic Acid; DPA: Docosapentaenoic Acid; ALA: α-Linolenic Acid; EPA: Eicosapentaenoic Acid; DHA: Docosahexaenoic Acid; PUFA: PolyUnsaturated Fatty Acids; SFA: Saturated Fatty Acids; MUFA: MonoUnsaturated Fatty Acids.

A Sankey plot based on the literature was constructed (**Figure 4**) to highlight the associations of the candidate oxylipins (in color) with the main underlying processes involved in MetS, namely blood clotting, vascular tone, inflammation, endothelial integrity, glucose homeostasis and adipogenesis. All processes were represented supporting the potential of the oxylipin signature to provide a fine and integrative phenotyping of the MetS molecular pathways. Inflammation was the most enriched biological process with sixteen linked oxylipins followed by vascular tone (linked to eleven oxylipins) and glucose homeostasis (linked to ten oxylipins). Blood clotting and endothelial integrity are both linked to seven oxylipins. Adipogenesis was less well represented, being only linked to two candidate oxylipins. Eight candidate oxylipins have no function described so far.

**Figure 4.**
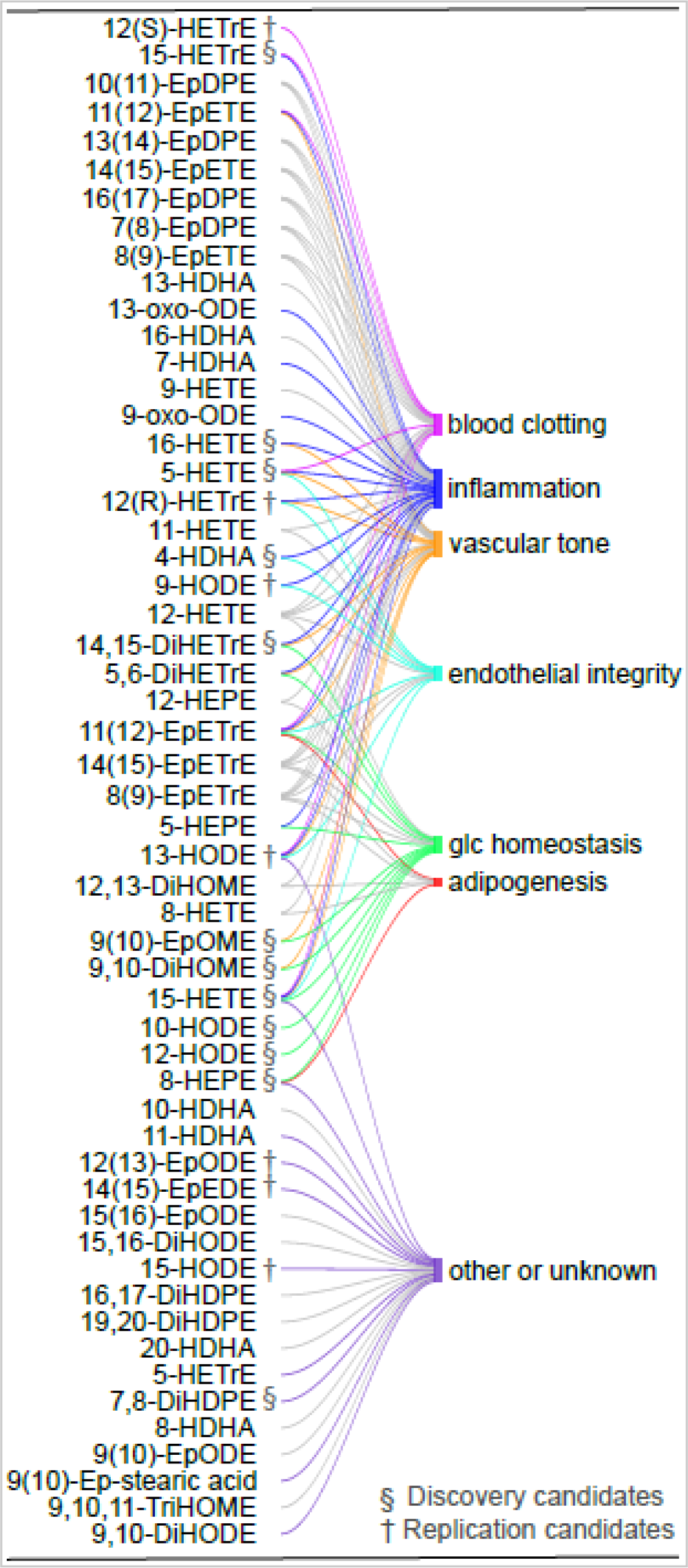
Biological functions of the candidate oxylipins discriminating participants with MetS. Sankey plot showing the relationships between the 54 oxylipins of the Discovery and Replication studies and the biological functions related to MetS (i.e. inflammation, vascular tone, blood clotting, endothelial permeability, adipogenesis and glucose homeostasis). The oxylipins selected in the Discovery and Replication studies (n=29) have their links highlighted in color. Relationships were established based on a systematic manual literature search including 110 references (∼2 studies/oxylipin). For 47% of these studies, experiments were realized using human in vitro models whereas 5·4% used in vivo human experiments and 27% were realized using in vivo animal experiments. The package R “stringr” was used to establish the links between the oxylipins and the cardiometabolic functions, then the package “googleVis” was used to generate the Sankey plot.

### 3.4. The identified candidate oxylipins allow the construction of a robust predictive model of MetS

By combining the candidate oxylipins selected in the Discovery and Replication studies (n =29 oxylipins), a Least Absolute Shrinkage and Selection Operator (LASSO)-penalized conditional logistic regression model selecting 23 of the 29 candidate oxylipins was constructed with the participants of the Discovery study. The predictive performance of this LASSO model reached a cross-validated Area Under the Receiver Operating Characteristic Curve (AUC_ROC_) of 89% (95% confidence interval (CI) 85-93%) in the Discovery study (**Figure 5a**) and a cross-validated AUC_ROC_ of 78% (95% CI 72-85%) in the Replication study (**Figure 5b**). The model based on the 23 candidate oxylipins and fitted using the Discovery participants (i.e. training set) correctly classified 83% of the Discovery participants. When the model was applied to the Replication study (i.e. the validation set), 72% of the participants were correctly classified. The Case and Control participants of the Replication study being slightly less contrasted than those in the Discovery study (see Volcano plot **Figure 1**); this may have contributed to the lower precision of classification rate in the Replication study. By a combined effect, the 23 oxylipins retained by the LASSO model all contribute to the estimation of MetS risk. An additional analysis of odd ratios in the 23-oxylipins based model (as represented in the Circos plot in **Figure 5c** **and Supplemental Table S2**) shows that the variation in concentration of some of the oxylipins had a significant individual impact on the model’s estimation of risk. Specifically, in this model, MetS risk was positively associated with 8-HEPE (OR = 1.837, 95% CI: 1.54- 2.19, p <0.001), 9(10)-epoxy-stearic acid (OR = 1.179, 95% CI: 1.10-1.26, p <0.001), 16-HETE (OR = 1.182, 95% CI: 1.11-1.26, p <0.001), and 12(13)-EpODE (OR = 1.092, 95% CI: 1.02-1.18, p = 0.021) and negatively associated with 5-HEPE (OR = 0.595, 95% CI: 0.50-0.70, p <0.001), 15-HETE (OR = 0.805, 95% CI: 0.74-0.88, p <0.001), 9(10)-EpOME (OR = 0.882, 95% CI: 0.81-0.96, p =0.007), 7,8-DiHDPE (OR = 0.865, 95% CI: 0.82-0.91, p <0.001), and 4-HDHA (OR = 0.805, 95% CI: 0.68-0.97, p = 0.030).

**Figure 5.**
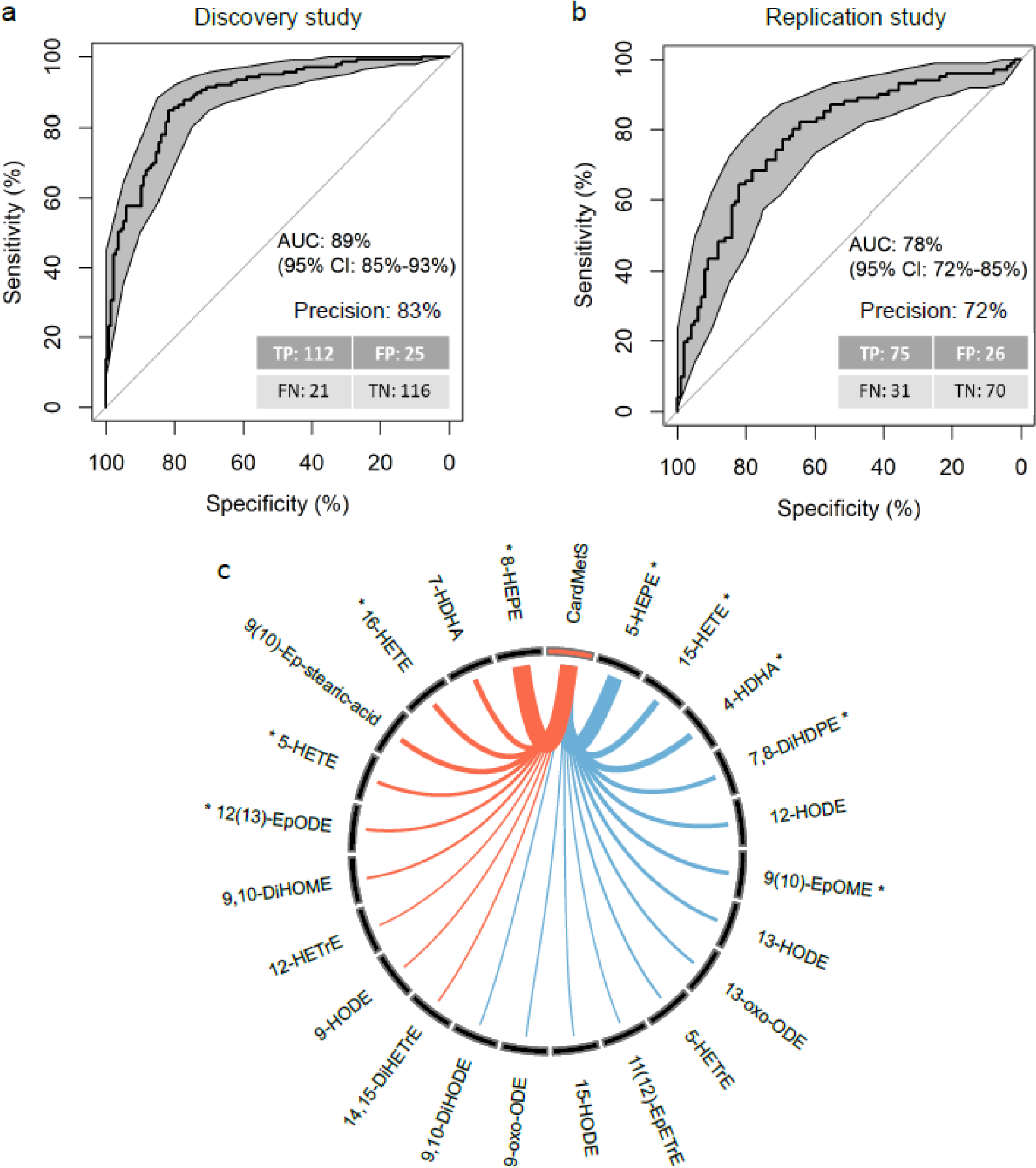
Optimization and external validation of the oxylipin signature of MetS. Based on the oxylipins selected in the Discovery and Replication cross-sectional studies (n=29 oxylipins), a LASSO-penalized conditional logistic regression model including 23 oxylipins was constructed with the participants of the Discovery study. The performances of the LASSO model were assessed in (**a**) the Discovery and (**b**) Replication cross-sectional studies using the Area Under the Receiver Operating Characteristic Curve (AUC and error rate were 10-fold cross-validated). TP: True Positive, FP: False Positive, TN: True Negative and FN: False Negative with Positive corresponding to CardMetS and Negative to Control. Circos plot (**c**) showing the odd ratios (OR) associated with each oxylipin selected in the LASSO model. * represent significant effect of oxylipins (p value <0·05) in the LASSO model. OR are for 1SD- increment of oxylipin absolute concentration. Line colors indicate if OR are >1 (orange) or <1 (blue). Line thickness represent the relative values of OR in the model.

### 3.5. The OxyScore derived from the LASSO predictive model is consistent and complementary with the criteria of MetS

The OxyScore (i.e. probability of having MetS according to the selected 23 candidate oxylipins) was computed for all participants of the Discovery study and correlated with the MetS-z-score (i.e. probability of having MetS according to the 5 criteria of MetS: waist circumference, triglycerides (TG), HDLc, blood pressure and fasting glucose) (**Figure 6**). The strength of the correlation informs on the consistency and the complementarity of the information brought by the OxyScore in comparison with the MetS criteria. The correlation between the OxyScore and the MetS-z-score was strong (r = 0.7) suggesting that a large part of the information provided by the OxyScore is consistent with the information provided by the MetS-z-score. However, the OxyScore also brings some information that is not captured by the MetS-z-score, if not the coefficient would be closer to 1.

**Figure 6.**
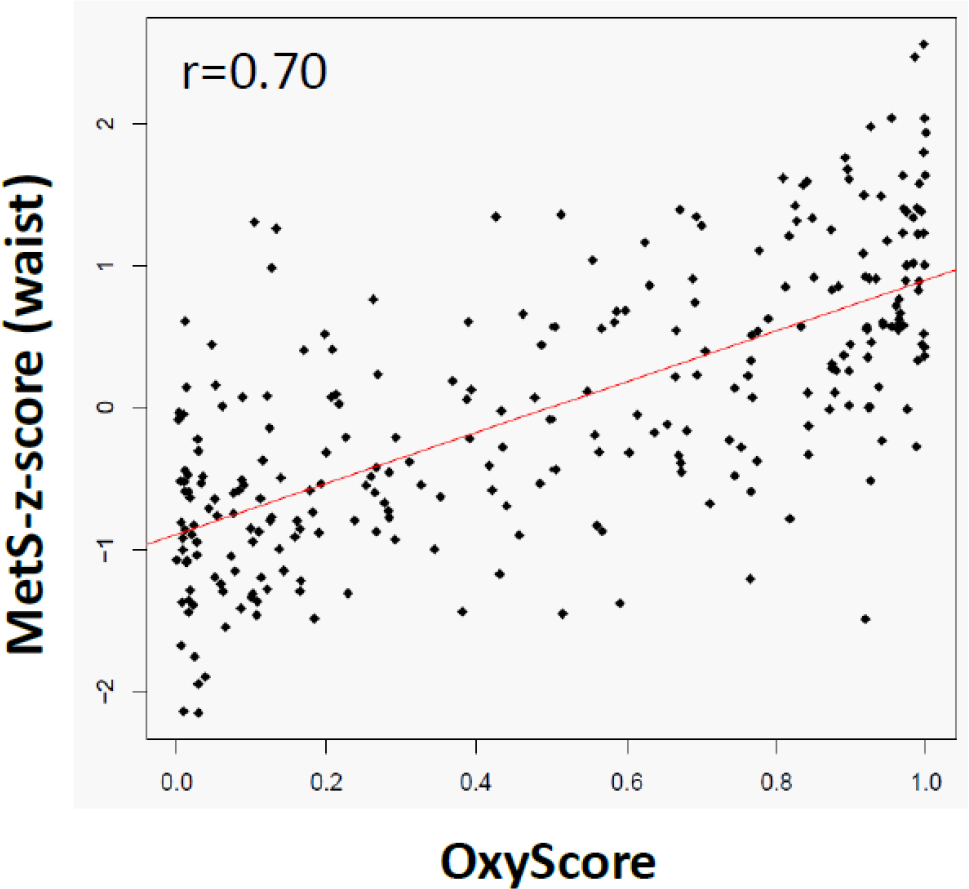
Relationships between the OxyScore and the Met-z-score. The OxyScore (i.e. probability of having MetS according to the identified and validated oxylipin signature, see Supp. Table 2) was computed for all participants of the Discovery cross-sectional study. Spearman correlation was established between the computed OxyScore and the Met-z-score. The Spearman correlation coefficient (r) was highly significant (p-value<0·001). The red line represents the linear orientation of the relation.

To go further in the assessment of the complementarity of the information provided by the oxylipin signature, we built a new LASSO model including the 5 MetS criteria additionally to the candidate oxylipins for the selection step (**Table 2**). All MetS criteria were selected in the new LASSO model consistently with the use of the MetS criteria as criteria of participant selection (see study design). More importantly, the new LASSO model also included most of the candidate oxylipins as only 6 candidate oxylipins (i.e. 9(10)-epoxy- stearic acid, 12-HETrE, 7-HDHA, 9,10-DiHOME, 9,10-DiHODE and 9(10)-EpOME) out of the 23 provided were not selected in the new LASSO model. This supports that most candidate oxylipins provide a characterization of MetS participants complementary to the MetS criteria.

**Table 2.** Odd ratios (OR) associated with the variable selected in the LASSO models constructed from the candidate oxylipins and/or the MetS criteria. Two new LASSO models were constructed with the participants of the Discovery study using either only the 5 criteria of MetS (i.e. waist circumference, blood pressure, fasting glucose, triglycerides and HDLc, LASSO model N°2) or the 5 criteria of MetS and the 23 candidate oxylipins (column 3, LASSO model N°3). Of note, the 5 MetS criteria were the criteria of selection of MetS participants. The predictive performance of these two LASSO models reached a cross-validated AUC of 93% (95% CI: 90-96%) and 95% (95% CI: 90-96%) respectively. Six candidate oxylipins (i.e. 9(10)-Ep-stearic acid, 12-HETrE, 7- HDHA, 9,10-DiHOME, 9,10-DiHODE and 9(10)-EpOME) out of the 23 provided were not selected in the LASSO model N°3.

|  | **Odd ratios associated with the variables selected in the LASSO models** |  |
| --- | --- | --- |
|  | **LASSO model N°2**<br>(using only the 5 MetS criteria) | **LASSO model N°3**<br>(using both the 23 candidate oxylipins and the 5 MetS criteria) |
| TG | 1.13 | 1.11 |
| SBP | 1.04 | 1.02 |
| DBP | 1.02 | 1.04 |
| Waist circumference | 1.18 | 1.13 |
| Fasting glucose | 1.15 | 1.10 |
| HDLc | 0.96 | 0.96 |
| 8-HEPE | -- | 1.53 |
| 16-HETE | -- | 1.16 |
| 12(13)-EpODE | -- | 1.01 |
| 5-HETE | -- | 0.11 |
| 9-HODE | -- | 1.01 |
| 14,15-DiHETrE | -- | 1.02 |
| 5-HETrE | -- | 0.95 |
| 15-HODE | -- | 0.96 |
| 13-oxo-ODE | -- | 0.93 |
| 11(12)-EpETrE | -- | 0.95 |
| 9-oxo-ODE | -- | 1.01 |
| 4-HDHA | -- | 0.92 |
| 13-HODE | -- | 0.95 |
| 12-HODE | -- | 0.91 |
| 7,8-DiHDPE | -- | 0.96 |
| 15-HETE | -- | 0.86 |
| 5-HEPE | -- | 0.71 |

In order to assess if the oxylipin signature was preferentially linked to one criterion of MetS, further correlation analysis and adjusted models were performed (see supplemental tables and figures). The correlation analysis show noticeable correlations between the OxyScore and waist circumference (r=0.57), fasting blood glucose (r=0.48), TG (r=0.60) and HDLc (r=0.51), but not with systolic and diastolic blood pressure (r=0.27) (**Supplemental Figure S3**). This may suggest that among the 23 candidate oxylipins used to predict the risk of having MetS, some of them provide information shared with waist circumference, fasting blood glucose, TG or HDLc. Correlations between each candidate oxylipins and each MetS criteria (**Supplemental Figure S4**) were mostly below 0.50, the strongest correlations (>0.30) being observed with the MetS-z-score and with TG: 12-HODE (−0.34 and -0.33 with MetS-z-score and TG respectively), 12,13- EpODE (0.41 with TG only) and 9(10)-epoxy-stearic acid (0.32 and 0.48 with MetS-z-score and TG respectively). Finally, when adjusting the LASSO model with each MetS criterion (one at a time), some oxylipins were no longer selected in the model (e.g. 12-HETrE, 7-HDHA and 9-HODE in the Waist- adjusted model, **see Supplemental Table S3**) suggesting that they were bringing similar information than the included MetS criterion (e.g. waist circumference). On the contrary, in the LASSO model adjusted with systolic or diastolic blood pressure, no oxylipin were excluded suggesting that the differences of blood pressure between Cases and Controls were not captured by the candidate oxylipins.

## 4. Discussion

Over the last decades, metabolomics and lipidomics have emerged as very relevant and powerful tools to capture the complexity of cardiometabolic disturbances and to inform on underlying mechanisms (6). Contrary to previous studies using data-driven approaches (8, 34), we have targeted a specific class of lipid mediators, namely oxylipins, which were hypothetized as being potential biologically relevant and integrative biomarkers of the molecular pathways of MetS. Our hypothesis-driven approach was combined with a high-throughput methodology allowing the quantitative profiling of over 130 different oxylipins. Through a rigorous process of selection and replication in two independent population studies involving 476 participants, we identified a panel of 29 oxylipins discriminant of MetS that provide a unique mechanistic signature and complementary information not revealed by the MetS clinical criteria. The model based on a refined signature of 23 oxylipins (coined as OxyScore) yielded excellent performances of classification and replicability.

In order to capture the full information brought by circulating oxylipins, we have analyzed total oxylipins (i.e. free and esterified). Integrating esterified oxylipins is important when investigating cardiometabolic health as they are found in lipoproteins (especially epoxy- and hydroxy-PUFA) (35, 36) that are involved in many cardiometabolic processes such as inflammation, oxidative stress or endothelial activation. Moreover, esterified oxylipins represent the major pool of circulating oxylipins. These may also arise from the secretion of blood cells (i.e. immune cells and platelets) or endothelium and they circulate unbound (free) or bound to plasma proteins such as albumin (37, 38). Other potential vectors of circulating oxylipins in plasma include extracellular vesicles that are both carriers and producers of oxylipins (39, 40).

The biological interpretation of circulating oxylipins requires considering them both as biomarkers and actors of biological processes. In the Discovery study, the oxylipin signature highlighted lower levels of linoleic acid (LA) derived oxylipins (i.e. HODEs including 9- and 13-HODE and their oxo-derivatives as well as 10- and 12-HODE) in the MetS group. These 18 carbon oxylipins can arise from the LOX pathways and/or from free-radical mediated oxidation (41). In terms of biological activities, 9- and 13-HODE have been described as potent regulators of monocytes/macrophages and neutrophils in which they stimulate lipid uptake (42, 43) and induce apoptosis (44). Concerning the inflammatory response, 13-HODE inhibits the production of the chemoattractant LTB_4_ by isolated human neutrophils (45, 46) while 9-HODE induces the expression of the pro-inflammatory mediators TNFα and MIP-2α in RAW264.7 macrophages (47). Both HODEs were also showed to prevent the activation of endothelial cells (48–51); to decrease the secretion of the fibrinolytic inhibitor plasminogen activator inhibitor-1 (PAI-1) (52) and to inhibit platelet aggregation (46, 53, 54). Neither the biological activities nor the formation route of 10- and 12-HODE have been precisely described in humans but high levels of 10- and 12-HODE have been associated with insulin resistance (33). Our results are consistent with a previous clinical trial showing lower levels of 9- and 13- HODE in very low density lipoproteins (VLDL) and in LDL from MetS subjects (n=17) in comparison with healthy controls (n=14) (55). Another small clinical trial reported higher levels of 9- and 13-HODE in LDL of MetS participants in comparison with healthy controls (56). However, the technical approach to quantify HODEs (i.e. thin-layer chromatography *vs* liquid chromatography coupled with tandem mass spectrometry (LC-MS/MS)) and the absence of antioxidant during sample preparation may have induced artificial production of HODEs (19). Altogether, the low levels of HODEs in the oxylipin signature of the MetS participants of the Discovery study reflects a decreased LOX activity and/or a well-controlled systemic oxidative stress probably linked to the activation of the antioxidant systems. Considering the important regulatory functions of 9- and 13-HODEs, their low levels in MetS participants may indicate that inflammation, endothelial and platelet functions were under control.

The oxylipin signature of MetS was also characterized by high levels of epoxy-PUFAs and low levels of vicinal dihydroxy-PUFAs (i.e. metabolites of epoxy-PUFAs produced by the sEH) reflecting an activation of the CYP pathway and a reduced activity of sEH. Notably, in the Discovery study, MetS participants exhibited higher levels of 12(13)-EpODE, 14(15)-EpEDE and 9(10)-epoxy-stearic acid while 9,10- DiHOME and 7,8-DiHDPE were significantly lower. In general, epoxy-PUFAs are reported to be protective in regards to cardiometabolic disease while activation of sEH is usually associated with metabolic stress (57). CYP enzymes are highly expressed in the liver but they are also found in the kidney, heart, skeletal muscle, pancreas, adipose tissue, endothelium, leucocytes and platelets (58, 59). So far, the biological activities of epoxy-PUFAs have been mainly reported for those derived from AA such as 11(12)-EpETrE. This oxylipin has important roles in the regulation of cardiometabolic health as it has been showed to improve glucose homeostasis (60–67) and endothelial function (68–72), to induce vasodilatation (72–79), to modulate platelet aggregation (80–82) and adipogenesis (63, 78, 83) and to inhibit leucocyte adhesion to the vascular wall (84). The effects of 11(12)-EpETE derived from EPA and 9(10)EPOME derived from LA are also partly documented: the former was shown to inhibit platelet aggregation (82) and induce vasodilatation (85) while the latter enhanced insulin signaling in HepG2 cells (86). To the best of our knowledge, the biological activities of 12(13)-EpODE and 14(15)-EpEDE have not been investigated so far. High levels of 9(10)-epoxy-stearic acid may induce lipid accumulation and oxidative stress in HepG2 cells (87, 88). Among the vicinal dihydroxy-PUFAs that were substantially and significantly reduced in MetS participants, 9,10-DiHOME was shown to increase coronary resistance (89) and to attenuate insulin signaling (86) but no information is available for 7,8-DiHDPE. Vicinal dihydroxy-PUFAs derived from AA (i.e. 5,6-DiHETrE and 14,15-DiHETrE) have been shown to facilitate chemoattraction of monocytes (90), to induce vasorelaxation in canine microcirculation and mouse arteries (73, 74) but less potently than their corresponding epoxide, and to attenuate insulin signaling (86). Changes in the CYP:sEH axis in the favor to epoxy-PUFAs in MetS participants might seem counterintuitive, but it was already reported in a clinical trial comparing healthy volunteers to patients with coronary artery disease (CAD) associated or not with obesity (91). The higher Epoxy-PUFAs:Diols ratios in CAD patients was suggested to be a compensatory response to the presence of advanced cardiovascular disease. Similarly, it can be hypothesized that MetS participants have activated CYP and decreased sEH activity to compensate for their cardiometabolic disturbances. In a clinical trial comparing Controls vs MetS participants, this compensatory mechanism was not observed (92). However, it is worth mentioning that age was a major confounding factor in this study and the significantly lower mean age of the Control participants (32 vs 51 y) could have contributed to the higher epoxide levels independently of the absence of MetS (93).

Finally, the oxylipin signature of the MetS participants in the Discovery study also suggested modification of the metabolism of long chain PUFAs (i.e. ≥ 20 carbons) by LOX enzymes. Concerning the eicosanoids (i.e. oxylipins derived from the 20 carbons PUFAs), the 5-LOX products of AA and EPA (i.e. 5-HETE and 5-HEPE) were lower while 15-LOX products (15-HETrE and 15-HETE) were higher in MetS. Interestingly, these differences support the hypothesis of a compensatory mechanism since 5-LOX products are rather detrimental to cardiometabolic health *via* the induction of inflammation (94–97), vasoconstriction (98), endothelial dysfunction (98) and platelet aggregation (99) while 15-LOX products have resolving action by reducing polymorphonuclear leukocyte (PMN) activation (100) and chemotaxis (45, 94) possibly *via* negative feedback regulation of 5-LOX (101). Effects of MetS on the 5-LOX products of DHA (i.e. 4- HDHA and 7-HDHA) were inconsistent with one found higher and the other lower. Of note, the biological activities of these oxylipins remain poorly described except for their binding capacity to peroxisome proliferator-activated receptor gamma (PPARγ) (102, 103) and a potential antiangiogenic effect of 4-HDHA (104). Similarly, the lower amount of the DGLA product 12-HETrE is difficult to interpret, as its metabolic origin is unclear (from platelet 12-LOX or CYP hydroxylase (105)) and depending on its configuration (S or R) it can be respectively anti-thrombotic (106, 107) or pro-inflammatory (108).

Although it might seem counterintuitive, the oxylipin signature of the MetS participants of the Discovery study may reflect the implementation of compensatory mechanisms including (i) the control of oxidative stress, (ii) the modulation of the CYP:sEH axis in favor of the protective epoxy-PUFAs and (iii) the production of regulatory oxylipins such as the 15-LOX products of long chain PUFAs to negatively regulate the 5-LOX pathway.

The oxylipin signature of MetS identified in the Discovery study was highly replicable in the Replication study. To the best of our knowledge, no metabolomic or lipidomic signature of the MetS has been externally validated so far (6). Moreover, the differences of oxylipin concentrations between Cases and Controls were mostly similar in both studies. Basically, in the Replication study, we also observed modulations of the CYP:sEH axis indicating a higher Epoxide:Diol ratio in MetS participants as a potential compensatory mechanism for the cardiometabolic disturbances. However, in the Replication study, the levels of 9- and 13-HODE were not lower in the MetS participants but rather tended to be higher. Another difference concerns the 5-LOX products of DGLA, AA and EPA (i.e. 5-HETrE, 5-HETE and 5-HEPE) that became significantly higher suggesting an active perturbation of cardiometabolic health associated with inflammation, vasoconstriction, endothelial dysfunction and platelet aggregation. Finally, the pathway involving the CYP ω-hydroxylase remains perturbed in the Replication study but in the opposite direction and more profoundly with 15-HODE levels being substantially lower while 16-HETE became significantly higher. The biological function of 15-HODE is unknown but 16-HETE has been described as an anti-inflammatory mediator *via* the inhibition of human PMN adhesion and aggregation and of LTB_4_ synthesis (109). The high levels 9- and 13-HODE associated with the higher production of 15- and 16-HETE could reflect the implementation of compensatory mechanisms. However, contrary to what was observed in the Discovery study, it does not seem sufficient to counterbalance the activation of the detrimental 5-LOX pathway. Of note, this pathway is known to be activated with aging (110) and MetS participants of the Replication study were significantly older than those of the Discovery study. This could have contributed to this impaired control of the 5-LOX pathway and may indicate an increased risk of cardiometabolic diseases.

We acknowledge limitations of our study. A first potential limitation concerns the sample size of the studies that does not allow stratifying the analysis for example to look at the results among younger and older participants. Moreover, because of discrepancies in the oxylipin coverage of both MS platforms (88 vs 58 oxylipins), we performed the selection of oxylipins using a smaller matrix including the oxylipins quantified in both platforms (54 oxylipins). However, it should be noted that this 54 oxylipin matrix covers all classes of oxylipins. The identified oxylipin signature of MetS requires further validation in various and larger-scale populations to confirm and extend the findings of this first investigation.

## 5. Conclusion

From a clinical perspective, the oxylipin signature of MetS we have identified not only have excellent performances of classification and replicability but most importantly, it provides a unique and integrative characterization of molecular pathways of MetS. This could allow a better understanding and stratification of the risk of cardiometabolic diseases. More precisely, the originality of the oxylipin signature relies on the information it provides on several key molecular pathways of MetS including oxidative stress, inflammation and the regulation of vascular tone, blood clotting, endothelial permeability, glucose homeostasis and adipogenesis. This level of information is not provided by the MetS clinical criteria (i.e. WC, BP, TG, HDLc, glycaemia) and it could help to distinguish patients with similar clinical diagnosis (i.e. MetS or not based on the 5 MetS criteria) but with a different level of risk of cardiometabolic diseases. In the Discovery and Replication studies, the oxylipin signature suggested the implementation of compensatory mechanism of the cardiometabolic disturbances in the MetS participants that could be impaired with age. The oxylipin signature of MetS is also original with regards to another MetS signature recently identified using untargeted metabolomics (8). Indeed, this approach does not allow the detection of oxylipins and rather informs on the most abundant circulating metabolites. In their study, Comte and collaborators identified a 26 metabolites- signature having excellent performances of classification (AUC of the PLS-DA model: 0.96, 95% CI: 0.94- 0.99) but it was not validated in an independent study. This metabolic signature provides information on various metabolic pathways (i.e. the urea cycle, amino-acid, sphingo and glycerophospholipid, and sugar metabolisms) different from those covered by the oxylipin signature.

## Conflict of interest

The authors have declared that no conflict of interest exists.

## Supporting information

Supplemental methods and results

Supplemental missing data

## Data Availability

All data produced in the present study are available upon reasonable request to the corresponding author

## Acknowledgements

The authors would like to thank Dominique Bayle, Séverine Valéro, Nicole Hartung, Nadja Kampschulte, Elisabeth Koch, Laura Kutzner and Katharina Rund for supporting sample preparation. This study was supported by French, German, Danish and Polish national research agencies (respectively ANR, grant N° ANR-16-HDHL-0004-01/02; the German Federal Ministry for Education and Research (BMBF), grant N°01EA1702; the Danish Innovation Fonden, grant N° 4203 – 00006B; and the Polish national center for research and development, grant N° ERAH.E220.18.001) in the framework of the Oxygenate project supported by the European Joint Programming Initiative “A Healthy Diet for a Healthy Life” (JPI HDHL; http://www.healthydietforhealthylife.eu/). Independent support was provided by USDA CRIS Project 2032-51530-025-00D to JWN. The USDA is an equal opportunity employer and provider.

The authors thank Younes Esseddik, Thi Hong Van Duong, Régis Gatibelza, Jagatjit Mohinder and Aladi Timera (computer scientists); Fabien Szabo de Edelenyi, Julien Allegre, Nathalie Arnault, Laurent Bourhis, Nicolas Dechamp (data-manager/statisticians); Cédric Agaësse, Alexandre De Sa, Rebecca Lutchia (dietitians), Merveille Kouam (health event validator); Maria Gomes (Nutrinaute support); and Nathalie Druesne-Pecollo (operational coordination) for their technical contribution to the NutriNet-Santé study. We thank all the volunteers of the NutriNet-Santé cohort. The NutriNet-Santé study was supported by the following public institutions: Ministère de la Santé, Santé Publique France, Institut National de la Santé et de la Recherche Médicale (INSERM), Institut National de Recherche pour l’Agriculture, l’Alimentation et l’Environnement (INRAE), Conservatoire National des Arts et Métiers (CNAM) and Université Sorbonne Paris Nord.

## Abbreviations

AA: arachidonic acid
AHEI: alternative healthy eating index
ALA: α-linolenic acid
AUC_ROC_: area under the receiver operating characteristic curves
BH: Benjamini-Hochberg
BHT: butylated hydroxytoluene
BMI: body mass index
BP: blood pressure
CAD: coronary artery disease
COX: cyclooxygenase
CYP: cytochrome P450
DGLA: dihomo-γ-linolenic acid
DHA: docosahexaenoic acid
EDTA: Ethylenediaminetetraacetic acid
EPA: eicosapentaenoic acid
FAMEs: fatty acids methyl esters
FFQ: food frequency questionnaire
GC: gas chromatography
HDLc: high density lipoprotein cholesterol
LA: linoleic acid
LASSO: least absolute shrinkage and selection operator
LC: liquid chromatography
LDLc: low density lipoprotein cholesterol
LOX: lipoxygenase
MeOH: methanol
MetS: metabolic syndrome
MRM: multiple reaction monitoring
MS: mass spectrometry
MS/MS: tandem mass spectrometry
OA: oleic acid
OR: odd ratio
PMN: polymorphonuclear
PPARγ: proliferator-activated receptor γ
PUFAs: polyunsaturated fatty acids
PURE: Prospective Urban and Rural Epidemiological
QC: quality control
SD: standard deviation
sEH: soluble epoxide hydrolase
SPE: solid phase extraction
TG: triglycerides
VLDL: very density lipoprotein cholesterol
WC: waist circumference.

## Footnotes

### Author Contributions

CG, JWN, MT, NHS and AS designed the study. CG, CD, LL, MT, MD and ABR performed participant selection. MT, LL, MD, AS, KZ and ABR extracted and sent samples and metadata. MM, NHS, JBM, JDC, CD and PCC prepared and analyzed the samples. MFH, AA, MM and NHS were in charge of data management and sharing. CD pre-processed the data. JT and CD performed statistical analysis under the supervision of MP and CG and with the help of MT, LL, MD, JWN, SLA and CS. CG and JWN drafted the manuscript. All authors contributed to writing, providing critical revision and all approved the submitted version of the manuscript.

CD and JT were two early career investigators (PhD student and postdoc) who equally contributed. CD is the first co-authors as the results presented here are part of her thesis work.

## Notes

**Funding sources** National research agencies: (i) French (grant N° ANR-16-HDHL-0004-01/02), (ii) German (BMBF grant N°01EA1702), (iii) Danish (grant N° 4203 – 00006B) and (iv) Polish (grant N° ERAH.E220.18.001) in the framework of the Oxygenate project supported by the European Joint Programming Initiative “A Healthy Diet for a Healthy Life” (JPI HDHL). And USDA CRIS Project 2032-51530-025-00D.

### Competing Interest Statement

The authors have declared no competing interest.

### Funding Statement

This study was funded by national research agencies: French grant ANR.16.HDHL.0004.01/02, German BMBF grant 01EA1702, Danish grant 4203.00006B and Polish grant ERAH.E220.18.001 in the framework of the Oxygenate project supported by the European Joint Programming Initiative A Healthy Diet for a Healthy Life (JPI HDHL). And USDA CRIS Project 2032.51530.025.00D.

### Author Declarations

PURE study was approved by the Institutional Review Board of the Wroclaw Medical University (IRB number: KB-443/2006). NutriNet-Sante study was approved by the International Research Board of the French Institute of Health and Medical research (IRB Inserm nb 0000388FWA00005831) and the Comite National Informatique et Liberte (CNIL 908450 and 909216).

## References

1. Saklayen, M. G. (2018) The Global Epidemic of the Metabolic Syndrome. Curr Hypertens Rep. 20(2):12.

2. Grundy, S. M. (2016) Metabolic syndrome update. Trends Cardiovasc Med. 26(4):364–73.

3. Alberti, K. G., Eckel, R. H., Grundy, S. M., Zimmet, P. Z., Cleeman, J. I., Donato, K. A., et al. (2009) Harmonizing the metabolic syndrome: a joint interim statement of the International Diabetes Federation Task Force on Epidemiology and Prevention; National Heart, Lung, and Blood Institute; American Heart Association; World Heart Federation; International Atherosclerosis Society; and International Association for the Study of Obesity. Circulation. 120(16):1640–5.

4. Robberecht, H., Hermans, N. (2016) Biomarkers of Metabolic Syndrome: Biochemical Background and Clinical Significance. Metab Syndr Relat Disord. 14(2):47–93.

5. O’Neill, S., Bohl, M., Gregersen, S., Hermansen, K., O’Driscoll, L. (2016) Blood-Based Biomarkers for Metabolic Syndrome. Trends Endocrinol Metab. 27(6):363–74.

6. Monnerie, S., Comte, B., Ziegler, D., Morais, J. A., Pujos-Guillot, E., Gaudreau, P. (2020) Metabolomic and Lipidomic Signatures of Metabolic Syndrome and its Physiological Components in Adults: A Systematic Review. Sci Rep. 10(1):669.

7. Gallart-Ayala, H., Teav, T., Ivanisevic, J. (2020) Metabolomics meets lipidomics: Assessing the small molecule component of metabolism. BioEssays. 42(12):2000052.

8. Comte, B., Monnerie, S., Brandolini-Bunlon, M., Canlet, C., Castelli, F., Chu-Van, E., et al. (2021) Multiplatform metabolomics for an integrative exploration of metabolic syndrome in older men. EBioMedicine. 69:103440.

9. Dennis, E. A., Norris, P. C. (2015) Eicosanoid storm in infection and inflammation. Nat Rev Immunol. 15(8):511–23.

10. van Ommen, B., Keijer, J., Heil, S. G., Kaput, J. (2009) Challenging homeostasis to define biomarkers for nutrition related health. Molecular Nutrition & Food Research. 53(7):795–804.

11. Fernández-García, J. C., Cardona, F., Tinahones, F. J. (2013) Inflammation, oxidative stress and metabolic syndrome: dietary modulation. Curr Vasc Pharmacol. 11(6):906–19.

12. Shearer, G. C., Newman, J. W. (2009) Impact of circulating esterified eicosanoids and other oxylipins on endothelial function. Curr Atheroscler Rep. 11(6):403–10.

13. Tourdot, B. E., Ahmed, I., Holinstat, M. (2014) The emerging role of oxylipins in thrombosis and diabetes. Front Pharmacol. 4:176.

14. Lopategi, A., López-Vicario, C., Alcaraz-Quiles, J., García-Alonso, V., Rius, B., Titos, E., et al. (2016) Role of bioactive lipid mediators in obese adipose tissue inflammation and endocrine dysfunction. Mol Cell Endocrinol. 419:44–59.

15. Mainka, M., Dalle, C., Pétéra, M., Dalloux-Chioccioli, J., Kampschulte, N., Ostermann, A. I., et al. (2020) Harmonized procedures lead to comparable quantification of total oxylipins across laboratories. J Lipid Res. 61(11):1424–36.

16. Rund, K. M., Ostermann, A. I., Kutzner, L., Galano, J. M., Oger, C., Vigor, C., et al. (2018) Development of an LC-ESI(-)-MS/MS method for the simultaneous quantification of 35 isoprostanes and isofurans derived from the major n3- and n6-PUFAs. Anal Chim Acta. 1037:63–74.

17. Teo, K., Chow, C. K., Vaz, M., Rangarajan, S., Yusuf, S. (2009) The Prospective Urban Rural Epidemiology (PURE) study: examining the impact of societal influences on chronic noncommunicable diseases in low-, middle-, and high-income countries. Am Heart J. 158(1):1–7.e1.

18. Hercberg, S., Castetbon, K., Czernichow, S., Malon, A., Mejean, C., Kesse, E., et al. (2010) The Nutrinet-Santé Study: a web-based prospective study on the relationship between nutrition and health and determinants of dietary patterns and nutritional status. BMC Public Health. 10(1):242.

19. Koch, E., Mainka, M., Dalle, C., Ostermann, A. I., Rund, K. M., Kutzner, L., et al. (2020) Stability of oxylipins during plasma generation and long-term storage. Talanta. 217:121074.

20. Ostermann, A. I., Koch, E., Rund, K. M., Kutzner, L., Mainka, M., Schebb, N. H. (2020) Targeting esterified oxylipins by LC–MS - Effect of sample preparation on oxylipin pattern. Prostaglandins & Other Lipid Mediators. 146:106384.

21. Kutzner, L., Rund, K. M., Ostermann, A. I., Hartung, N. M., Galano, J. M., Balas, L., et al. (2019) Development of an Optimized LC-MS Method for the Detection of Specialized Pro-Resolving Mediators in Biological Samples. Front Pharmacol. 10:169.

22. Lillington, J. M., Trafford, D. J. H., Makin, H. L. J. (1981) A rapid and simple method for the esterification of fatty acids and steroid carboxylic acids prior to gas-liquid chromatography. Clinica Chimica Acta. 111(1):91–8.

23. Chiuve, S. E., Fung, T. T., Rimm, E. B., Hu, F. B., McCullough, M. L., Wang, M., et al. (2012) Alternative dietary indices both strongly predict risk of chronic disease. J Nutr. 142(6):1009–18.

24. Gurka, M. J., Lilly, C. L., Oliver, M. N., DeBoer, M. D. (2014) An examination of sex and racial/ethnic differences in the metabolic syndrome among adults: a confirmatory factor analysis and a resulting continuous severity score. Metabolism. 63(2):218–25.

25. Zou, H., Hastie, T. (2005) Regularization and variable selection via the elastic net. Journal of the Royal Statistical Society: Series B (Statistical Methodology). 67(2):301–20.

26. Goeman, J. J. (2010) L1 penalized estimation in the Cox proportional hazards model. Biom J. 52(1):70–84.

27. Friedman, J. H., Hastie, T., Tibshirani, R. (2010) Regularization Paths for Generalized Linear Models via Coordinate Descent. 2010. 33(1):1–22.

28. Robin, X., Turck, N., Hainard, A., Tiberti, N., Lisacek, F., Sanchez, J. C., et al. (2011) pROC: an open-source package for R and S+ to analyze and compare ROC curves. BMC Bioinformatics. 12:77.

29. Tibshirani, R. J., Taylor, J., Lockhart, R., Tibshirani, R. (2016) Exact Post-Selection Inference for Sequential Regression Procedures. Journal of the American Statistical Association. 111(514):600–20.

30. Krzywinski, M., Schein, J., Birol, I., Connors, J., Gascoyne, R., Horsman, D., et al. (2009) Circos: an information aesthetic for comparative genomics. Genome Res. 19(9):1639–45.

31. Vangaveti, V. N., Jansen, H., Kennedy, R. L., Malabu, U. H. (2016) Hydroxyoctadecadienoic acids: Oxidised derivatives of linoleic acid and their role in inflammation associated with metabolic syndrome and cancer. Eur J Pharmacol. 785:70–6.

32. Yeung, J., Holinstat, M. (2017) Who is the real 12-HETrE? Prostaglandins Other Lipid Mediat. 132:25–30.

33. Umeno, A., Shichiri, M., Ishida, N., Hashimoto, Y., Abe, K., Kataoka, M., et al. (2013) Singlet oxygen induced products of linoleates, 10- and 12-(Z,E)-hydroxyoctadecadienoic acids (HODE), can be potential biomarkers for early detection of type 2 diabetes. PLoS One. 8(5):e63542.

34. Yin, X., Willinger, C. M., Keefe, J., Liu, J., Fernández-Ortiz, A., Ibáñez, B., et al. (2020) Lipidomic profiling identifies signatures of metabolic risk. EBioMedicine. 51:102520.

35. Schebb, N. H., Ostermann, A. I., Yang, J., Hammock, B. D., Hahn, A., Schuchardt, J. P. (2014) Comparison of the effects of long-chain omega-3 fatty acid supplementation on plasma levels of free and esterified oxylipins. Prostaglandins Other Lipid Mediat. 113–115:21-9.

36. Shearer, G. C., Newman, J. W. (2008) Lipoprotein lipase releases esterified oxylipins from very low-density lipoproteins. Prostaglandins Leukot Essent Fatty Acids. 79(6):215–22.

37. Ek-Von Mentzer, B. A., Zhang, F., Hamilton, J. A. (2001) Binding of 13-HODE and 15-HETE to phospholipid bilayers, albumin, and intracellular fatty acid binding proteins. implications for transmembrane and intracellular transport and for protection from lipid peroxidation. J Biol Chem. 276(19):15575–80.

38. Maclouf, J., Kindahl, H., Granström, E., Samuelsson, B. (1980) Interactions of prostaglandin H2 and thromboxane A2 with human serum albumin. Eur J Biochem. 109(2):561–6.

39. Boilard, E. (2018) Extracellular vesicles and their content in bioactive lipid mediators: more than a sack of microRNA. J Lipid Res. 59(11):2037–46.

40. Pizzinat, N., Ong-Meang, V., Bourgailh-Tortosa, F., Blanzat, M., Perquis, L., Cussac, D., et al. (2020) Extracellular vesicles of MSCs and cardiomyoblasts are vehicles for lipid mediators. Biochimie. 178:69–80.

41. Vangaveti, V., Baune, B. T., Kennedy, R. L. (2010) Hydroxyoctadecadienoic acids: novel regulators of macrophage differentiation and atherogenesis. Ther Adv Endocrinol Metab. 1(2):51–60.

42. Nagy, L., Tontonoz, P., Alvarez, J. G., Chen, H., Evans, R. M. (1998) Oxidized LDL regulates macrophage gene expression through ligand activation of PPARgamma. Cell. 93(2):229–40.

43. Fu, Y., Luo, N., Lopes-Virella, M. F., Garvey, W. T. (2002) The adipocyte lipid binding protein (ALBP/aP2) gene facilitates foam cell formation in human THP-1 macrophages. Atherosclerosis. 165(2):259–69.

44. Vangaveti, V. N., Shashidhar, V. M., Rush, C., Malabu, U. H., Rasalam, R. R., Collier, F., et al. (2014) Hydroxyoctadecadienoic acids regulate apoptosis in human THP-1 cells in a PPARγ-dependent manner. Lipids. 49(12):1181–92.

45. Iversen, L., Fogh, K., Bojesen, G., Kragballe, K. (1991) Linoleic acid and dihomogammalinolenic acid inhibit leukotriene B4 formation and stimulate the formation of their 15-lipoxygenase products by human neutrophils in vitro. Evidence of formation of antiinflammatory compounds. Agents Actions. 33(3-4):286–91.

46. Camp, R. D., Fincham, N. J. (1985) Inhibition of ionophore-stimulated leukotriene B4 production in human leucocytes by monohydroxy fatty acids. Br J Pharmacol. 85(4):837–41.

47. Warner, D. R., Liu, H., Miller, M. E., Ramsden, C. E., Gao, B., Feldstein, A. E., et al. (2017) Dietary Linoleic Acid and Its Oxidized Metabolites Exacerbate Liver Injury Caused by Ethanol via Induction of Hepatic Proinflammatory Response in Mice. Am J Pathol. 187(10):2232–45.

48. Delerive, P., Furman, C., Teissier, E., Fruchart, J., Duriez, P., Staels, B. (2000) Oxidized phospholipids activate PPARalpha in a phospholipase A2-dependent manner. FEBS Lett. 471(1):34–8.

49. Buchanan, M. R., Bertomeu, M. C., Haas, T. A., Orr, F. W., Eltringham-Smith, L. L. (1993) Localization of 13-hydroxyoctadecadienoic acid and the vitronectin receptor in human endothelial cells and endothelial cell/platelet interactions in vitro. Blood. 81(12):3303–12.

50. Haas, T. A., Bastida, E., Nakamura, K., Hullin, F., Admirall, L., Buchanan, M. R. (1988) Binding of 13-HODE and 5-, 12- and 15-HETE to endothelial cells and subsequent platelet, neutrophil and tumor cell adhesion. Biochim Biophys Acta. 961(2):153–9.

51. Haas, T. A., Bertomeu, M. C., Bastida, E., Buchanan, M. R. (1990) Cyclic AMP regulation of endothelial cell triacylglycerol turnover, 13-hydroxyoctadecadienoic acid (13-HODE) synthesis and endothelial cell thrombogenicity. Biochim Biophys Acta. 1051(2):174–8.

52. Marx, N., Bourcier, T., Sukhova, G. K., Libby, P., Plutzky, J. (1999) PPARgamma activation in human endothelial cells increases plasminogen activator inhibitor type-1 expression: PPARgamma as a potential mediator in vascular disease. Arterioscler Thromb Vasc Biol. 19(3):546–51.

53. Coene, M. C., Bult, H., Claeys, M., Laekeman, G. M., Herman, A. G. (1986) Inhibition of rabbit platelet activation by lipoxygenase products of arachidonic and linoleic acid. Thromb Res. 42(2):205–14.

54. Setty, B. N., Berger, M., Stuart, M. J. (1987) 13-Hydroxyoctadecadienoic acid (13-HODE) stimulates prostacyclin production by endothelial cells. Biochem Biophys Res Commun. 146(2):502–9.

55. Shearer, G. C., Borkowski, K., Puumala, S. L., Harris, W. S., Pedersen, T. L., Newman, J. W. (2018) Abnormal lipoprotein oxylipins in metabolic syndrome and partial correction by omega-3 fatty acids. Prostaglandins Leukot Essent Fatty Acids. 128:1–10.

56. Colas, R., Pruneta-Deloche, V., Guichardant, M., Luquain-Costaz, C., Cugnet-Anceau, C., Moret, M., et al. (2010) Increased lipid peroxidation in LDL from type-2 diabetic patients. Lipids. 45(8):723–31.

57. Dos Santos, L. R. B., Fleming, I. (2020) Role of cytochrome P450-derived, polyunsaturated fatty acid mediators in diabetes and the metabolic syndrome. Prostaglandins Other Lipid Mediat. 148:106407.

58. Anzenbacher, P., Hudecek, J. (2001) Differences in flexibility of active sites of cytochromes P450 probed by resonance Raman and UV-Vis absorption spectroscopy. J Inorg Biochem. 87(4):209–13.

59. Zhu, Y., Schieber, E. B., McGiff, J. C., Balazy, M. (1995) Identification of arachidonate P-450 metabolites in human platelet phospholipids. Hypertension. 25(4 Pt 2):854-9.

60. Falck, J. R., Manna, S., Moltz, J., Chacos, N., Capdevila, J. (1983) Epoxyeicosatrienoic acids stimulate glucagon and insulin release from isolated rat pancreatic islets. Biochem Biophys Res Commun. 114(2):743–9.

61. Huang, H., Weng, J., Wang, M. H. (2016) EETs/sEH in diabetes and obesity-induced cardiovascular diseases. Prostaglandins Other Lipid Mediat. 125:80–9.

62. Sonnweber, T., Pizzini, A., Nairz, M., Weiss, G., Tancevski, I. (2018) Arachidonic Acid Metabolites in Cardiovascular and Metabolic Diseases. Int J Mol Sci. 19(11).

63. Xu, H., Fu, J. L., Miao, Y. F., Wang, C. J., Han, Q. F., Li, S., et al. (2016) Prostaglandin E2 receptor EP3 regulates both adipogenesis and lipolysis in mouse white adipose tissue. J Mol Cell Biol. 8(6):518–29.

64. Ma, B., Xiong, X., Chen, C., Li, H., Xu, X., Li, X., et al. (2013) Cardiac-specific overexpression of CYP2J2 attenuates diabetic cardiomyopathy in male streptozotocin-induced diabetic mice. Endocrinology. 154(8):2843–56.

65. Chen, C., Wang, D. W. (2013) CYP epoxygenase derived EETs: from cardiovascular protection to human cancer therapy. Curr Top Med Chem. 13(12):1454–69.

66. Luo, P., Chang, H. H., Zhou, Y., Zhang, S., Hwang, S. H., Morisseau, C., et al. (2010) Inhibition or deletion of soluble epoxide hydrolase prevents hyperglycemia, promotes insulin secretion, and reduces islet apoptosis. J Pharmacol Exp Ther. 334(2):430–8.

67. Aliwarga, T., Evangelista, E. A., Sotoodehnia, N., Lemaitre, R. N., Totah, R. A. (2018) Regulation of CYP2J2 and EET Levels in Cardiac Disease and Diabetes. Int J Mol Sci. 19(7).

68. Bellien, J., Iacob, M., Remy-Jouet, I., Lucas, D., Monteil, C., Gutierrez, L., et al. (2012) Epoxyeicosatrienoic acids contribute with altered nitric oxide and endothelin-1 pathways to conduit artery endothelial dysfunction in essential hypertension. Circulation. 125(10):1266–75.

69. Imig, J. D. (2016) Epoxyeicosatrienoic Acids and 20-Hydroxyeicosatetraenoic Acid on Endothelial and Vascular Function. Adv Pharmacol. 77:105–41.

70. Imig, J. D. (2019) Epigenetic soluble epoxide hydrolase regulation causes endothelial dysfunction. Acta Physiol (Oxf). 225(1):e13203.

71. Yang, L., Cheriyan, J., Gutterman, D. D., Mayer, R. J., Ament, Z., Griffin, J. L., et al. (2017) Mechanisms of Vascular Dysfunction in COPD and Effects of a Novel Soluble Epoxide Hydrolase Inhibitor in Smokers. Chest. 151(3):555–63.

72. Hamzaoui, M., Guerrot, D., Djerada, Z., Duflot, T., Richard, V., Bellien, J. (2018) Cardiovascular consequences of chronic kidney disease, impact of modulation of epoxyeicosatrienoic acids. Ann Cardiol Angeiol (Paris). 67(3):141–8.

73. Hercule, H. C., Schunck, W. H., Gross, V., Seringer, J., Leung, F. P., Weldon, S. M., et al. (2009) Interaction between P450 eicosanoids and nitric oxide in the control of arterial tone in mice. Arterioscler Thromb Vasc Biol. 29(1):54–60.

74. Oltman, C. L., Weintraub, N. L., VanRollins, M., Dellsperger, K. C. (1998) Epoxyeicosatrienoic acids and dihydroxyeicosatrienoic acids are potent vasodilators in the canine coronary microcirculation. Circ Res. 83(9):932–9.

75. Chabowski, D. S., Cohen, K. E., Abu-Hatoum, O., Gutterman, D. D., Freed, J. K. (2020) Crossing signals: bioactive lipids in the microvasculature. Am J Physiol Heart Circ Physiol. 318(5):H1185–h97.

76. Khan, A. H., Falck, J. R., Manthati, V. L., Campbell, W. B., Imig, J. D. (2014) Epoxyeicosatrienoic acid analog attenuates angiotensin II hypertension and kidney injury. Front Pharmacol. 5:216.

77. Roman, R. J. (2002) P-450 metabolites of arachidonic acid in the control of cardiovascular function. Physiol Rev. 82(1):131–85.

78. Romashko, M., Schragenheim, J., Abraham, N. G., McClung, J. A. (2016) Epoxyeicosatrienoic Acid as Therapy for Diabetic and Ischemic Cardiomyopathy. Trends Pharmacol Sci. 37(11):945–62.

79. Proctor, K. G., Falck, J. R., Capdevila, J. (1987) Intestinal vasodilation by epoxyeicosatrienoic acids: arachidonic acid metabolites produced by a cytochrome P450 monooxygenase. Circ Res. 60(1):50–9.

80. Krötz, F., Riexinger, T., Buerkle, M. A., Nithipatikom, K., Gloe, T., Sohn, H. Y., et al. (2004) Membrane-potential-dependent inhibition of platelet adhesion to endothelial cells by epoxyeicosatrienoic acids. Arterioscler Thromb Vasc Biol. 24(3):595–600.

81. Fitzpatrick, F. A., Ennis, M. D., Baze, M. E., Wynalda, M. A., McGee, J. E., Liggett, W. F. (1986) Inhibition of cyclooxygenase activity and platelet aggregation by epoxyeicosatrienoic acids. Influence of stereochemistry. J Biol Chem. 261(32):15334–8.

82. VanRollins, M. (1995) Epoxygenase metabolites of docosahexaenoic and eicosapentaenoic acids inhibit platelet aggregation at concentrations below those affecting thromboxane synthesis. J Pharmacol Exp Ther. 274(2):798–804.

83. Li, R., Xu, X., Chen, C., Wang, Y., Gruzdev, A., Zeldin, D. C., et al. (2015) CYP2J2 attenuates metabolic dysfunction in diabetic mice by reducing hepatic inflammation via the PPARγ. Am J Physiol Endocrinol Metab. 308(4):E270–82.

84. Node, K., Huo, Y., Ruan, X., Yang, B., Spiecker, M., Ley, K., et al. (1999) Anti-inflammatory properties of cytochrome P450 epoxygenase-derived eicosanoids. Science. 285(5431):1276–9.

85. Zhang, Y., Oltman, C. L., Lu, T., Lee, H. C., Dellsperger, K. C., VanRollins, M. (2001) EET homologs potently dilate coronary microvessels and activate BK(Ca) channels. Am J Physiol Heart Circ Physiol. 280(6):H2430–40.

86. Bettaieb, A., Nagata, N., AbouBechara, D., Chahed, S., Morisseau, C., Hammock, B. D., et al. (2013) Soluble epoxide hydrolase deficiency or inhibition attenuates diet-induced endoplasmic reticulum stress in liver and adipose tissue. J Biol Chem. 288(20):14189–99.

87. Liu, Y., Cheng, Y., Li, J., Wang, Y., Liu, Y. (2018) Epoxy Stearic Acid, an Oxidative Product Derived from Oleic Acid, Induces Cytotoxicity, Oxidative Stress, and Apoptosis in HepG2 Cells. J Agric Food Chem. 66(20):5237–46.

88. Liu, Y., Li, J., Liu, Y. (2020) Effects of epoxy stearic acid on lipid metabolism in HepG2 cells. J Food Sci. 85(10):3644–52.

89. Edin, M. L., Wang, Z., Bradbury, J. A., Graves, J. P., Lih, F. B., DeGraff, L. M., et al. (2011) Endothelial expression of human cytochrome P450 epoxygenase CYP2C8 increases susceptibility to ischemia-reperfusion injury in isolated mouse heart. Faseb j. 25(10):3436–47.

90. Kundu, S., Roome, T., Bhattacharjee, A., Carnevale, K. A., Yakubenko, V. P., Zhang, R., et al. (2013) Metabolic products of soluble epoxide hydrolase are essential for monocyte chemotaxis to MCP-1 in vitro and in vivo. J Lipid Res. 54(2):436–47.

91. Theken, K. N., Schuck, R. N., Edin, M. L., Tran, B., Ellis, K., Bass, A., et al. (2012) Evaluation of cytochrome P450-derived eicosanoids in humans with stable atherosclerotic cardiovascular disease. Atherosclerosis. 222(2):530–6.

92. Ramirez, C. E., Shuey, M. M., Milne, G. L., Gilbert, K., Hui, N., Yu, C., et al. (2014) Arg287Gln variant of EPHX2 and epoxyeicosatrienoic acids are associated with insulin sensitivity in humans. Prostaglandins Other Lipid Mediat. 113–115:38-44.

93. Mitchell, S. J., Kane, A. E., Hilmer, S. N. (2011) Age-related changes in the hepatic pharmacology and toxicology of paracetamol. Curr Gerontol Geriatr Res. 2011:624156.

94. Nayeem, M. A. (2018) Role of oxylipins in cardiovascular diseases. Acta Pharmacol Sin. 39(7):1142–54.

95. Goetzl, E. J., Pickett, W. C. (1980) The human PMN leukocyte chemotactic activity of complex hydroxy-eicosatetraenoic acids (HETEs). J Immunol. 125(4):1789–91.

96. Valone, F. H., Franklin, M., Sun, F. F., Goetzl, E. J. (1980) Alveolar macrophage lipoxygenase products of arachidonic acid: isolation and recognition as the predominant constituents of the neutrophil chemotactic activity elaborated by alveolar macrophages. Cell Immunol. 54(2):390–401.

97. Stenson, W. F., Parker, C. W. (1980) Monohydroxyeicosatetraenoic acids (HETEs) induce degranulation of human neutrophils. J Immunol. 124(5):2100–4.

98. Maayah, Z. H., El-Kadi, A. O. (2016) The role of mid-chain hydroxyeicosatetraenoic acids in the pathogenesis of hypertension and cardiac hypertrophy. Arch Toxicol. 90(1):119–36.

99. Setty, B. N., Werner, M. H., Hannun, Y. A., Stuart, M. J. (1992) 15-Hydroxyeicosatetraenoic acid-mediated potentiation of thrombin-induced platelet functions occurs via enhanced production of phosphoinositide-derived second messengers--sn-1,2-diacylglycerol and inositol-1,4,5-trisphosphate. Blood. 80(11):2765–73.

100. Smith, R. J., Justen, J. M., Nidy, E. G., Sam, L. M., Bleasdale, J. E. (1993) Transmembrane signaling in human polymorphonuclear neutrophils: 15(S)-hydroxy-(5Z,8Z,11Z,13E)-eicosatetraenoic acid modulates receptor agonist-triggered cell activation. Proc Natl Acad Sci U S A. 90(15):7270–4.

101. Petrich, K., Ludwig, P., Kühn, H., Schewe, T. (1996) The suppression of 5-lipoxygenation of arachidonic acid in human polymorphonuclear leucocytes by the 15-lipoxygenase product (15S)-hydroxy- (5Z,8Z,11Z,13E)-eicosatetraenoic acid: structure-activity relationship and mechanism of action. Biochem J. 314:911–6.

102. Chen, K. M., Thompson, H., Vanden-Heuvel, J. P., Sun, Y. W., Trushin, N., Aliaga, C., et al. (2021) Lipoxygenase catalyzed metabolites derived from docosahexaenoic acid are promising antitumor agents against breast cancer. Sci Rep. 11(1):410.

103. González-Périz, A., Planagumà, A., Gronert, K., Miquel, R., López-Parra, M., Titos, E., et al. (2006) Docosahexaenoic acid (DHA) blunts liver injury by conversion to protective lipid mediators: protectin D1 and 17S-hydroxy-DHA. Faseb j. 20(14):2537–9.

104. Sapieha, P., Stahl, A., Chen, J., Seaward, M. R., Willett, K. L., Krah, N. M., et al. (2011) 5-Lipoxygenase metabolite 4-HDHA is a mediator of the antiangiogenic effect of ω-3 polyunsaturated fatty acids. Sci Transl Med. 3(69):69ra12.

105. Yeung, J., Hawley, M., Holinstat, M. (2017) The expansive role of oxylipins on platelet biology. J Mol Med (Berl). 95(6):575–88.

106. Ikei, K. N., Yeung, J., Apopa, P. L., Ceja, J., Vesci, J., Holman, T. R., et al. (2012) Investigations of human platelet-type 12-lipoxygenase: role of lipoxygenase products in platelet activation. J Lipid Res. 53(12):2546–59.

107. Yeung, J., Tourdot, B. E., Adili, R., Green, A. R., Freedman, C. J., Fernandez-Perez, P., et al. (2016) 12(S)-HETrE, a 12-Lipoxygenase Oxylipin of Dihomo-γ-Linolenic Acid, Inhibits Thrombosis via Gαs Signaling in Platelets. Arterioscler Thromb Vasc Biol. 36(10):2068–77.

108. Masferrer, J. L., Rimarachin, J. A., Gerritsen, M. E., Falck, J. R., Yadagiri, P., Dunn, M. W., et al. (1991) 12(R)-hydroxyeicosatrienoic acid, a potent chemotactic and angiogenic factor produced by the cornea. Exp Eye Res. 52(4):417–24.

109. Bednar, M. M., Gross, C. E., Russell, S. R., Fuller, S. P., Ahern, T. P., Howard, D. B., et al. (2000) 16(R)-hydroxyeicosatetraenoic acid, a novel cytochrome P450 product of arachidonic acid, suppresses activation of human polymorphonuclear leukocyte and reduces intracranial pressure in a rabbit model of thromboembolic stroke. Neurosurgery. 47(6):1410–8; discussion 8-9.

110. Caligiuri, S. P., Aukema, H. M., Ravandi, A., Pierce, G. N. (2014) Elevated levels of pro-inflammatory oxylipins in older subjects are normalized by flaxseed consumption. Exp Gerontol. 59:51–7.

