## Supplemental methods and results for "Oxylipin profiling identifies a mechanistic signature of metabolic syndrome: results from two independent cohorts"

### Supplemental Method

#### Procedure of MS data preprocessing

- **MS data integration and imputation**

The profiling of oxylipins used is based on Mainka et al. (1) and allows the detection of 133 oxylipins. During MS data integration (MultiQuant version 2.1.1 (Sciex) or MassHunter Quantitative Analysis version B.09.00 (Agilent), depending on the MS platform running the MS analysis), peaks with low analytical quality preventing precise quantification were flagged as follows (**Table 1**):

***Table 1. Flags corresponding to peak characteristics***

| Flags | Peak characteristics |
|---|---|
| **Flag 1** | <LLOQ[1] |
| **Flag 2** | >ULOQ[2] |
| **Flag 3** | Double peak |
| **Flag 4** | Peak shoulder |
| **Flag 5** | Peak shape |
| **Flag 6** | Coelution matrix |
| **Flag 7** | No integration possible |

[1]LLOD: lower limit of detection

[2]ULOQ: upper limit of quantification

Peaks flagged as 1, 6 and 7 were considered as missing data. Only peaks flagged 1 and 7 were imputed (see below) whereas flag 6 were replaced by NA. For peaks flagged as 2, the automatic value of ULOQ was kept in the dataset. For peaks flagged as 3, 4 and 5, the manually integrated data was kept in the dataset.

Before missing data imputation, the percentage of missing data was assessed study by study, and group by group. If an oxylipin presented more than 30% of missing data, this oxylipin was excluded of the matrix. The **figure 1** represents the strategy and the number of oxylipins excluded. The distribution and percentage of missing data for retained oxylipins are shown in **supplemental excel file**.

Missing data (flags 1 and 7) were imputed as follows. For flag 1, the imputed value consisted in a random value automatically chosen between the limit of detection (LOD) and the lower limit of quantification (LLOQ). Indeed, flag 1 indicated peaks that were successfully detected but in an amount being not high enough to be accurately quantified. This choice of imputation enables to keep the information of an existing low quantity detected. For flag 7, the imputed value consisted in a random value automatically chosen between a value representing 10% of the LOD and the LOD. Indeed, for these recorded signals there is no means to determine whether the corresponding compounds were absent from the sample or in too low abundance to be detected. The chosen imputation method enables to keep the information of a very low quantity without claiming an absolute absence of compound.

After exclusion of oxylipins having >30% of missing data (i.e. flag 1, 6 and 7) and missing data imputation, the final oxylipin matrix for the different studies were as showed in the **Figure 1** below:

(**a**) Discovery cross-sectional study

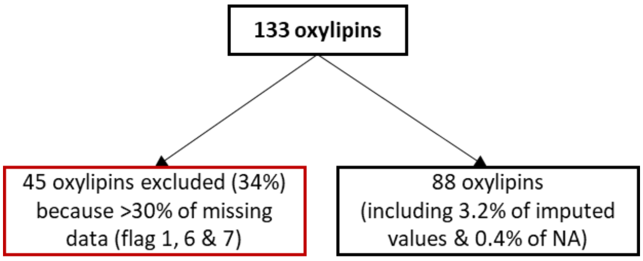


(**b**) Replication cross-sectional study

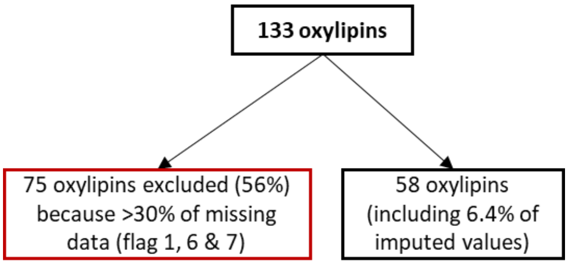


***Figure 1. Strategy and final oxylipin matrix for the Discovery (a) and Replication (b) cross-sectional studies***

- **MS data normalization (signal drift correction)**

MS analysis of large number of samples usually requires signal drift correction of data. For that, quality controls (i.e. a pool constituted of 5µL of each sample extract) were injected every 15 or 20 samples (depending on the MS platform running the MS analysis) to monitor stability of the analytical system and allow signal drift and batch effect correction. Since oxylipin profiling is a quantitative method based on the use of internal standards, the signal drift was very limited in our datasets and did not required signal drift correction except for a subset of samples. Briefly, among the 274 PURE samples analyzed, 50 had to be reinjected which is known to affect specific class of oxylipins (1). By using Galaxy web-based platform Workflow4Metabolomics (2), we corrected this signal drift according to the algorithm described by Van der Kloet et al. (3). Even if no re-injection occurs during the analysis of the Nutrinet-Santé samples, the same signal drift correct process was applied to ensure comparability of datasets.

- **Data adjustment**

To reduce the impact of total oxylipin levels on data variability, discrete intensity adjustments were performed. Each sample was assigned to a global concentration class based on its total oxylipin value (classes from A to L (depending on the study), defined by log2 intervals of 1, **Table 2**).

***Table 2. Concentration class according to the total oxylipin values study by study***

| Study | Total oxylipin value based on | Classes | Log2(total oxylipin) range |
|---|---|---|---|
| **Discovery study (PURE cohort)** | 88 oxylipins | A to H | x<12 to x>18 |
| **Replication study (NutriNet-Santé cohort)** | 58 oxylipins | A to L | x<10 to x>12 |

Using these intensity classes as homogeneous groups of samples, each oxylipin was individually adjusted. Firstly, each sample value of one oxylipin was reduced using its class mean value for the concerned oxylipin. All values were then multiplied by the population oxylipin mean (i.e. mean of all classes) to preserve the original scale of the concerned oxylipin. In other words, for each sample value of each oxylipin the following process was applied: ([sample value of oxylipin] / [sample class' mean of oxylipin])*([oxylipin population mean]). This per-oxylipin adjustment enabled adaptive normalisation, preventing disproportioned correction due to variable impact of total oxylipin effect on features. Of note, mean calculations were performed on each study group independently, to prevent biological effect erasure that could arise from unbalanced proportions in total oxylipin classes.

Of note, for the validation stage of the cross-sectional study, the oxylipin matrix as well as the oxylipin data adjustment had to be harmonized. As described previously, a common matrix of 54 oxylipins was used to discover and validate the candidate oxylipins and therefore the data adjustment was also performed on a total oxylipin value based on 54 oxylipins. This new data adjustment had no impact on oxylipin selection.

**Supplemental Tables & Figures**

*Supplemental Table S1. Plasma oxylipin and fatty acid absolute concentration for the Discovery and the Replication studies*

| | DISCOVERY study[1] | | | REPLICATION study[2] | | |
|---|---|---|---|---|---|---|
| **Variables** | **Controls (n=137) Mean (SD)** | **Cases (n=137) Mean (SD)** | **p-value[3]** | **Controls (n=101) Mean (SD)** | **Cases (n=101) Mean (SD)** | **p-value[3]** |
| | | | **Oxylipins (nM)** | | | |
| 9(10)-Ep stearic acid | 377.1 (118.3) | 452.8 (183.0) | <0.01 | 423.5 (130.6) | 477.3 (133.7) | <0.01 |
| 9-HODE | 14313.3 (2560.9) | 12892.7 (2785.5) | <0.001 | 269.3 (68.2) | 282.9 (84.2) | NS |
| 10-HODE | 56.1 (12.3) | 46.2 (12.3) | <0.001 | 14.4 (4.0) | 14.1 (4.6) | NS |
| 12-HODE | 152.4 (56.2) | 106.8 (40.8) | <0.001 | 8.5 (2.8) | 8.5 (3.1) | NS |
| 13-HODE | 29604.4 (6777.7) | 24226.4 (5799.4) | <0.001 | 350.0 (75.4) | 367.3 (73.6) | NS |
| 15-HODE | 8.4 (4.1) | 8.8 (5.7) | NS | 28.6 (22.8) | 21.7 (19.9) | <0.05 |
| 9-oxo-ODE | 8365.6 (4145.2) | 5599.7 (2528.6) | <0.001 | 47.8 (15.1) | 53.9 (18.3) | NS |
| 13-oxo-ODE | 14829.4 (6914.2) | 11059.8 (4641.7) | <0.001 | 542.6 (151.8) | 541.3 (174.5) | NS |
| 9(10)-EpOME | 351.7 (94.0) | 358.9 (126.7) | NS | 580.1 (121.8) | 573.3 (114.5) | NS |
| 9,10-DiHOME | 28.7 (11.0) | 24.8 (12.3) | <0.01 | 72.1 (29.5) | 64.1 (27.2) | NS |
| 12,13-DiHOME | 10.4 (4.2) | 8.5 (4.6) | <0.001 | 29.5 (13.2) | 25.7 (11.4) | NS |
| 9,10,11-TriHOME | 71.1 (46.3) | 47.4 (35.9) | <0.001 | 2.6 (1.8) | 2.5 (1.6) | NS |
| 9(10)-EpODE | 6.4 (3.4) | 9.2 (6.2) | <0.001 | 10.0 (5.6) | 11.4 (6.9) | NS |
| 12(13)-EpODE | 3.4 (1.6) | 4.9 (3.3) | <0.001 | 7.0 (4.0) | 7.6 (3.8) | NS |
| 15(16)-EpODE | 25.9 (15.2) | 40.1 (35.9) | <0.001 | 107.5 (75.2) | 119.8 (67.6) | NS |
| 9,10-DiHODE | 0.6 (0.3) | 0.6 (0.5) | NS | 2.4 (1.6) | 1.7 (0.8) | <0.01 |
| 15,16-DiHODE | 14.7 (8.3) | 14.7 (10.1) | NS | 68.8 (39.3) | 54.3 (25.8) | NS |
| 5-HETrE | 201.0 (100.0) | 231.3 (110.1) | NS | 3.3 (2.0) | 5.1 (3.0) | <0.001 |
| 12-HETrE | 2489.5 (948.3) | 2382.9 (766) | NS | 8.7 (6.0) | 9.9 (7.4) | NS |
| 15-HETrE | 1050.6 (411.9) | 1089.3 (368.9) | NS | 9.7 (3.7) | 11.3 (3.5) | <0.05 |
| 14(15)-EpEDE | 11.4 (4.3) | 13.7 (6.0) | <0.01 | 23.6 (7.7) | 27.7 (8.1) | <0.01 |
| 5-HETE | 3110.3 (1050.2) | 2980 (949.7) | NS | 54.3 (16.9) | 60.2 (15.7) | <0.05 |
| 8-HETE | 2150.9 (756.7) | 2257 (618.1) | NS | 40.5 (12.7) | 43.1 (12.2) | NS |
| 9-HETE | 3033.7 (687.6) | 3136 (632.8) | NS | 40.9 (27.5) | 42.2 (27.6) | NS |
| 11-HETE | 4751.7 (1469.7) | 4962.7 (1249.9) | NS | 54.4 (18.8) | 58.8 (16.9) | NS |
| 12-HETE | 3362.4 (1168.5) | 3608.1 (1020.8) | NS | 44.9 (16.1) | 48.0 (15.2) | NS |
| 15-HETE | 2357.8 (856.3) | 2401.2 (684.9) | NS | 51.4 (21.4) | 57.5 (21.2) | NS |
| 16-HETE | 18.9 (6.4) | 18.9 (6.2) | NS | 2.1 (0.8) | 2.5 (0.9) | <0.05 |
| 8(9)-EpETrE | 45.0 (17.4) | 50 (18.3) | NS | 85.4 (28.4) | 92.6 (30.2) | NS |
| 11(12)-EpETrE | 50.1 (20.6) | 54.4 (20.6) | NS | 129.8 (32.3) | 138.3 (34.4) | NS |

| | | | | | | |
|---|---|---|---|---|---|---|
| 14(15)-EpETrE | 75.9 (32.5) | 83.2 (31.3) | NS | 180.8 (46.2) | 192.9 (49.1) | NS |
| 5,6-DiHETrE | 21.9 (7.3) | 22.7 (8.2) | NS | 51.3 (14.5) | 57.5 (16.1) | <0.05 |
| 14,15-DiHETrE | 4.1 (1.2) | 3.9 (1.0) | NS | 3.5 (1.0) | 3.4 (0.8) | NS |
| 5-HEPE | 317.6 (182.0) | 302.9 (162) | NS | 10.1 (5.5) | 11.9 (5.2) | <0.05 |
| 8-HEPE | 221.7 (125.9) | 249.3 (126.9) | NS | 4.7 (3.3) | 5.0 (2.8) | NS |
| 12-HEPE | 559.5 (340.6) | 623.4 (331.1) | NS | 9.5 (6.2) | 9.8 (5.8) | NS |
| 8(9)-EpETE | 6.9 (4.3) | 8.1 (4.7) | NS | 17.3 (15) | 18.1 (13.1) | NS |
| 11(12)-EpETE | 5.1 (5.3) | 5.4 (4.8) | NS | 19.6 (12.4) | 20.4 (10.7) | NS |
| 14(15)-EpETE | 6.0 (4.3) | 6.4 (4.1) | NS | 24.2 (14.9) | 24.2 (13.4) | NS |
| 4-HDHA | 1313.9 (577.6) | 1172.9 (564.5) | NS | 24.2 (7.7) | 23.9 (7.6) | NS |
| 7-HDHA | 686.8 (353.1) | 718.8 (368.9) | NS | 15.4 (6.0) | 15.5 (5.6) | NS |
| 8-HDHA | 738.4 (316.2) | 737.5 (308) | NS | 15.0 (5.2) | 15.3 (5.9) | NS |
| 10-HDHA | 740.8 (345.8) | 758.4 (354.1) | NS | 14.7 (6.0) | 14.7 (5.8) | NS |
| 11-HDHA | 1419.9 (679.6) | 1475.5 (696.5) | NS | 20.5 (8.7) | 21.4 (8.3) | NS |
| 13-HDHA | 905.2 (416.6) | 902 (417.0) | NS | 18.2 (6.4) | 18.3 (6.8) | NS |
| 16-HDHA | 759.9 (340.3) | 754.1 (329.1) | NS | 15.5 (5.9) | 15.4 (5.2) | NS |
| 20-HDHA | 1114.0 (480.7) | 1060.9 (442.3) | NS | 28.2 (9.7) | 28.0 (9.4) | NS |
| 7(8)-EpDPE | 15.3 (6.7) | 15.9 (7.9) | NS | 40.3 (14.2) | 39.9 (15.4) | NS |
| 10(11)-EpDPE | 14.4 (6.2) | 15.9 (8.0) | NS | 40.9 (12.1) | 41.9 (14.3) | NS |
| 13(14)-EpDPE | 14.0 (6.5) | 14.8 (7.5) | NS | 38.9 (11.8) | 38.4 (12.7) | NS |
| 16(17)-EpDPE | 13.8 (6.4) | 14.4 (7.2) | NS | 34.3 (10.7) | 33.6 (12.3) | NS |
| 7,8-DiHDPE | 3.0 (1.9) | 2.3 (1.3) | <0.01 | 12.5 (5.0) | 10.6 (4.3) | <0.05 |
| 16,17-DiHDPE | 9.9 (4.2) | 10 (4.1) | NS | 2.4 (0.9) | 2.3 (0.7) | NS |
| 19,20-DiHDPE | 2.9 (1.4) | 2.4 (1.2) | <0.01 | 8.1 (6.0) | 7.3 (5.2) | NS |
| **Plasmatic Fatty acids (µg/mL)** | | | | | | |
| 14:0 | 25.6 (10.4) | 36.1 (22.0) | <0.001 | 10.5 (7.7) | 17.1 (16.5) | <0.01 |
| 16:0 | 711.1 (188.0) | 811.4 (294.9) | <0.05 | 513.1 (194.1) | 648.4 (326.2) | <0.001 |
| 16:1n-7 | 58.2 (24.4) | 79.6 (50.1) | <0.001 | 38.2 (17.6) | 61.9 (42.8) | <0.001 |
| 18:0 | 250.5 (64.2) | 271.2 (87.2) | NS | 229.1 (89.7) | 242.2 (61.5) | <0.05 |
| 18:1n-9 | 639.4 (178.4) | 784.0 (317.6) | <0.001 | 613.5 (174.9) | 746.2 (248.5) | <0.001 |
| 18:1n-7 | 52.9 (16.7) | 61.9 (25.5) | <0.05 | 53.9 (18.2) | 67.6 (33.5) | <0.001 |
| 18:2n-6 (LA) | 801.5 (265.1) | 781.3 (262.7) | NS | 832 (224.9) | 830.7 (259.8) | NS |
| 18:3n-6 (GLA) | 7.6 (4.6) | 10.7 (7.0) | <0.001 | 12.1 (8.7) | 15.1 (9.7) | <0.05 |
| 18:3n-3 (ALA) | 19.4 (9.1) | 24.6 (14.3) | <0.05 | 17.2 (10.0) | 20.4 (11.2) | <0.05 |
| 18 :4n-3 | n.d. | n.d. | n.d. | 6.0 (4.5) | 7.2 (5.1) | <0.05 |
| 22 :1n-9 | n.d. | n.d. | n.d. | 4.1 (1.5) | 4.3 (1.7) | NS |

| | | | | | | |
|---|---|---|---|---|---|---|
| 20:0 | 2.1 (1.5) | 2.4 (1.5) | NS | n.d. | n.d. | n.d. |
| 20:1n-9 | 6.7 (2.9) | 7.7 (3.8) | NS | n.d. | n.d. | n.d. |
| 20:2n-6 | 6.2 (2.2) | 6.5 (2.6) | NS | 9.2 (3.7) | 11.0 (4.0) | <0.05 |
| 20:3n-6 (DGLA) | 39.5 (16.4) | 45.7 (19.5) | <0.05 | 40.9 (14.7) | 48.4 (16.4) | <0.05 |
| 20:4n-6 (AA) | 179.4 (83.0) | 188.2 (77.0) | NS | 210.0 (54.0) | 230.1 (60.4) | NS |
| 22:0 | 0.8 (0.3) | 0.8 (0.4) | NS | n.d. | n.d. | n.d. |
| 22 :4n-6 | n.d. | n.d. | n.d. | 5.2 (2.8) | 5.9 (2.3) | <0.05 |
| 20:4n-3 | 3.7 (3.9) | 4.2 (3.2) | <0.05 | n.d. | n.d. | n.d. |
| 20:5n-3 (EPA) | 24.9 (22) | 25.8 (18.0) | NS | 43.8 (30.0) | 45.4 (21.3) | NS |
| 24:0 | 3.2 (1.8) | 3.4 (1.9) | NS | n.d. | n.d. | n.d. |
| 24:1n-9 | 1.2 (0.5) | 1.2 (0.6) | NS | n.d. | n.d. | n.d. |
| 22 :3n-3 | n.d. | n.d. | n.d. | 6.2 (3.9) | 6.4 (3.2) | NS |
| 22:5n-3 (DPA) | 12.8 (6.6) | 14.0 (7.4) | NS | 15.0 (6.0) | 16.4 (6.4) | NS |
| 22:6n-3 (DHA) | 52.9 (31.1) | 51.9 (27.9) | NS | 76.6 (28.2) | 76.4 (23.9) | NS |

Control for participants with <3 criteria and Case for participants with ≥3 criteria of cardiometabolic syndrome (obesity, high blood pressure, hypertriglyceridemia, low HDL-c and hyperglycemia).

n.d. for not determined.

[1]Selected participants were matched on gender, age (2y classes), smoking status (never+former vs current) and physical activity (low vs moderate+intense)

[2]Selected participants were matched on gender, age (2y classes), smoking status (never/former/irregular/current), physical activity (low/moderate/intense), menopausal status (NA/yes/no) and season of blood draw (winter/spring/summer/fall)

[3]Differences between Case and Control for the oxylipins and the plasmatic fatty acids were assed using univariate analysis (non-parametric Wilcoxon signed-rank test followed by Benjamini-Hochberg (BH) multiple tests correction) taking into consideration the matching of participants. NS for non-significant when the p-value is >0.05

*Supplemental Table S2. Oxylipins selected in the LASSO-penalized conditional logistic regression model*

Based on the LASSO-penalized conditional logistic regression model, the probability of being a Case (coined OxyScore) was calculated for each participant as follows:

OxyScore $= \frac{\exp(B*X)}{1+\exp(B*X)}$ where (B*X) corresponds to a combination of $b_0*x_0$, $b_1*x_1$, …, $b_n*x_n$ (n the number of oxylipins in the model) with $b_n$ being the adjusted coefficient of the n-th oxylipin in the optimized Lasso regression model and $x_n$ the corresponding oxylipin concentration. The 23 oxylipins included in the model and their respective coefficients, odd ratios, 95% confidence interval (CI) and p-value are in the table.

| Oxylipins included in the final/validated Lasso regression model | Coefficients | Odd ratios | 95% CI | p-value |
|---|---|---|---|---|
| 8-HEPE | 4.190 | 1.837 | (1.54-2.19) | <0.001 |
| 9(10)-Ep-stearic acid | 1.408 | 1.179 | (1.10-1.26) | <0.001 |
| 16-HETE | 1.246 | 1.182 | (1.11-1.26) | <0.001 |
| 12(13)-EpODE | 0.798 | 1.092 | (1.02-1.17) | 0.024 |
| 12-HETrE | 0.479 | 1.048 | (0.92-1.19) | 0.253 |
| 7-HDHA | 0.409 | 1.188 | (0.97-1.40) | 0.076 |
| 9,10-DiHOME | 0.365 | 1.075 | (0.99-1.15) | 0.076 |
| 5-HETE | 0.343 | 1.120 | (1.02-1.22) | 0.030 |
| 9-HODE | 0.263 | 1.045 | (0.93-1.12) | 0.264 |
| 14,15-DiHETrE | 0.006 | 1.018 | (0.81-1.08) | 0.622 |
| 9,10-DiHODE | -0.051 | 0.963 | (0.91-1.07) | 0.270 |
| 5-HETrE | -0.142 | 0.931 | (0.87-1.00) | 0.058 |
| 15-HODE | -0.296 | 0.956 | (0.91-1.03) | 0.138 |
| 13-oxo-ODE | -0.360 | 0.918 | (0.79-1.03) | 0.097 |
| 11(12)-EpETrE | -0.380 | 0.956 | (0.89-1.08) | 0.283 |
| 9-oxo-ODE | -0.634 | 0.959 | (0.87-1.24) | 0.475 |
| 4-HDHA | -0.775 | 0.805 | (0.68-0.97) | 0.030 |
| 13-HODE | -0.896 | 0.897 | (0.79-1.06) | 0.130 |
| 12-HODE | -0.948 | 0.874 | (0.78-1.00) | 0.051 |
| 7,8-DiHDPE | -0.999 | 0.865 | (0.82-0.91) | <0.001 |
| 9(10)-EpOME | -1.027 | 0.882 | (0.81-0.96) | 0.007 |
| 15-HETE | -1.302 | 0.805 | (0.74-0.88) | <0.001 |
| 5-HEPE | -3.683 | 0.595 | (0.50-0.70) | <0.001 |

***Supplemental Table S3. Impact of the addition of the MetS criteria in the Lasso model on the odd ratios and p-value of the 23 candidate oxylipins.***

Odd ratios for each of the 23 candidate oxylipins included in the Lasso model (created in the Discovery study and validated in the Replication study) adjusted with each individual cardiometabolic criteria (waist circumference, systolic blood pressure or SBP, diastolic blood pressure or DBP, fasting glucose, triglycerides and HDL). Empty cases mean that the oxylipin was excluded of the adjusted Lasso model.

| | **Odd ratios** | | | | | | |
|---|---|---|---|---|---|---|---|
| | **No adjust.** | **Waist** | **SBP** | **DBP** | **Fasting Glc** | **TG** | **HDL** |
| | **--** | **1.20** | **1.09** | **1.09** | **1.14** | **1.20** | **0.89** |
| 8-HEPE | 1.84 | 1.79 | 1.85 | 1.84 | 1.61 | 0.86 | 1.81 |
| 9(10)-Ep-stearic acid | 1.18 | 1.12 | 1.19 | 1.17 | 1.17 | | 1.12 |
| 16-HETE | 1.18 | 1.16 | 1.17 | 1.18 | 1.15 | 0.96 | 1.20 |
| 12(13)-EpODE | 1.09 | 1.07 | 1.09 | 1.09 | 1.06 | 0.91 | 1.06 |
| 12-HETrE | 1.05 | | 1.04 | 1.04 | 1.04 | 0.96 | 1.05 |
| 7-HDHA | 1.19 | | 1.15 | 1.14 | 1.18 | 0.95 | 1.13 |
| 9,10-DiHOME | 1.08 | 1.07 | 1.06 | 1.06 | | 1.01 | 1.07 |
| 5-HETE | 1.12 | 1.13 | 1.13 | 1.13 | 1.13 | 0.67 | 1.12 |
| 9-HODE | 1.05 | | 1.06 | 1.05 | 1.04 | 1.66 | 1.03 |
| 14,15-DiHETrE | 1.02 | 1.04 | 1.02 | 1.02 | 0.99 | 0.88 | 1.02 |
| 9,10-DiHODE | 0.96 | 0.97 | 0.97 | 0.97 | 0.98 | | 0.97 |
| 5-HETrE | 0.93 | 0.93 | 0.92 | 0.93 | 0.96 | | 0.93 |
| 15-HODE | 0.96 | 0.95 | 0.95 | 0.96 | 0.97 | 0.88 | 0.96 |
| 13-oxo-ODE | 0.92 | 0.92 | 0.92 | 0.91 | 0.96 | 0.90 | 0.92 |
| 11(12)-EpETrE | 0.96 | 0.96 | 0.95 | 0.95 | 0.92 | | 0.98 |
| 9-oxo-ODE | 0.96 | 0.99 | 0.98 | 0.99 | 0.96 | 1.04 | 0.94 |
| 4-HDHA | 0.81 | 0.93 | 0.82 | 0.83 | 0.80 | 1.21 | 0.83 |
| 13-HODE | 0.90 | 0.98 | 0.89 | 0.89 | 0.89 | 1.03 | 0.91 |
| 12-HODE | 0.87 | 0.85 | 0.88 | 0.88 | 0.91 | 1.05 | 0.90 |
| 7,8-DiHDPE | 0.87 | 0.94 | 0.88 | 0.88 | 0.90 | | 0.89 |
| 9(10)-EpOME | 0.88 | 0.90 | 0.87 | 0.87 | 0.94 | | 0.91 |
| 15-HETE | 0.81 | 0.82 | 0.81 | 0.81 | 0.83 | | 0.80 |
| 5-HEPE | 0.59 | 0.60 | 0.59 | 0.59 | 0.67 | 0.95 | 0.62 |

*Supplemental Figure S1. Participant's selection for the Discovery and Replication studies*

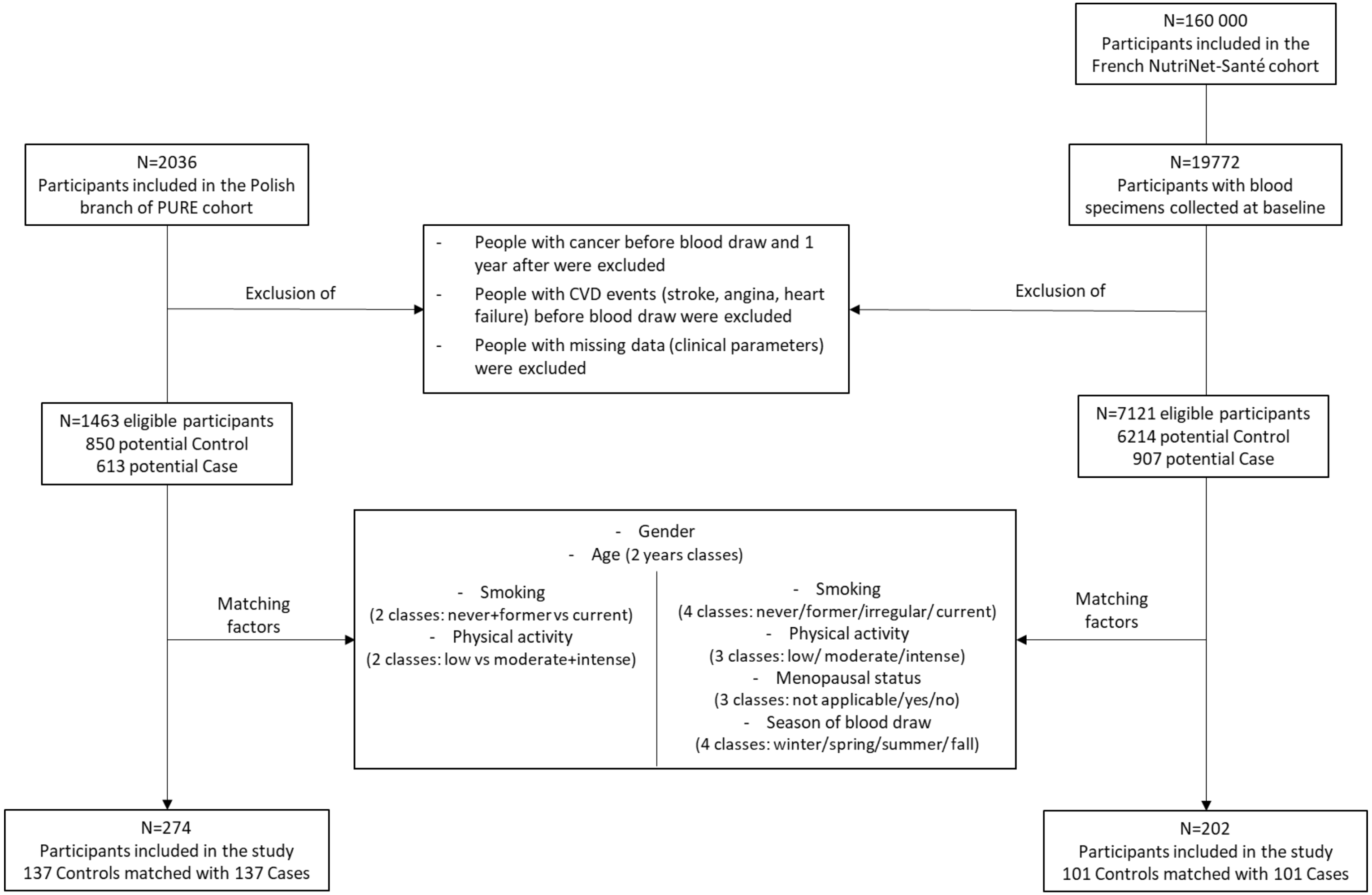


Control group: participants with <3 MetS criteria and Case group: participants with ≥3 MetS criteria (waist circumference, high blood pressure, hypertriglyceridemia, low HDL-c and hyperglycemia). To avoid the selection of incomparable Control participants between the two cohorts, the Control groups of each study were balanced according to the number of MetS criteria (i.e. 0, 1 or 2).

*Supplemental Figure S2. Selection of oxylipins discriminating the MetS in the Discovery and Replication studies*

Oxylipins were independently selected in (**a**) the Discovery study and (**b**) in the Replication study. Selection was done from a matrix of 54 common oxylipins using Elastic-Net penalized conditional logistic regression repeated on 350 bootstraps resampling. Oxylipins significantly associated with the odds of having MetS are ranked by decreasing frequency of selection across bootstraps. Oxylipins found in ≥80% of bootstraps (highlighted in blue) were selected. * Oxylipin put aside from subsequent analysis because of a putative low score of analytical robustness based previously published studies (1, 4) regarding (i) stability during transitory and long term storage, (ii) the technical and interlaboratory variabilities as well as the percentage of missing data imputation.

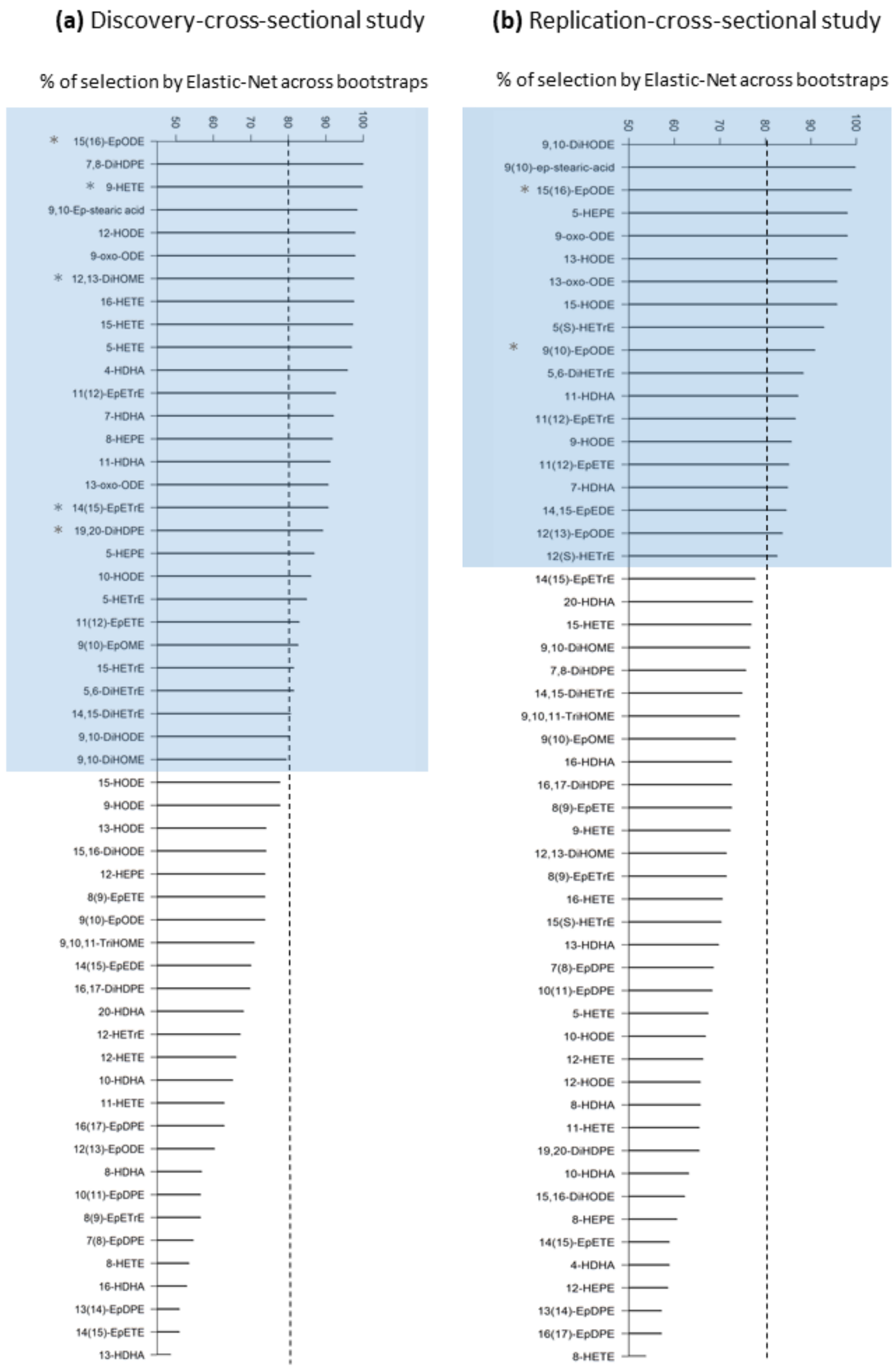

*Supplemental Figure S3. Relationships between the OxyScore and the criteria of MetS*

The OxyScore (i.e. probability of having MetS according to the identified and validated oxylipin signature, see Supp. Table 2) was computed for all participants of the Discovery cross-sectional study. Spearman correlations were established between the computed OxyScore and each individual criterion of the MetS. The Spearman correlation coefficient (r) were all highly significant (p-value<0.001) and were considered noteworthy when ≥0.5. The red line represents the linear orientation of the relation.

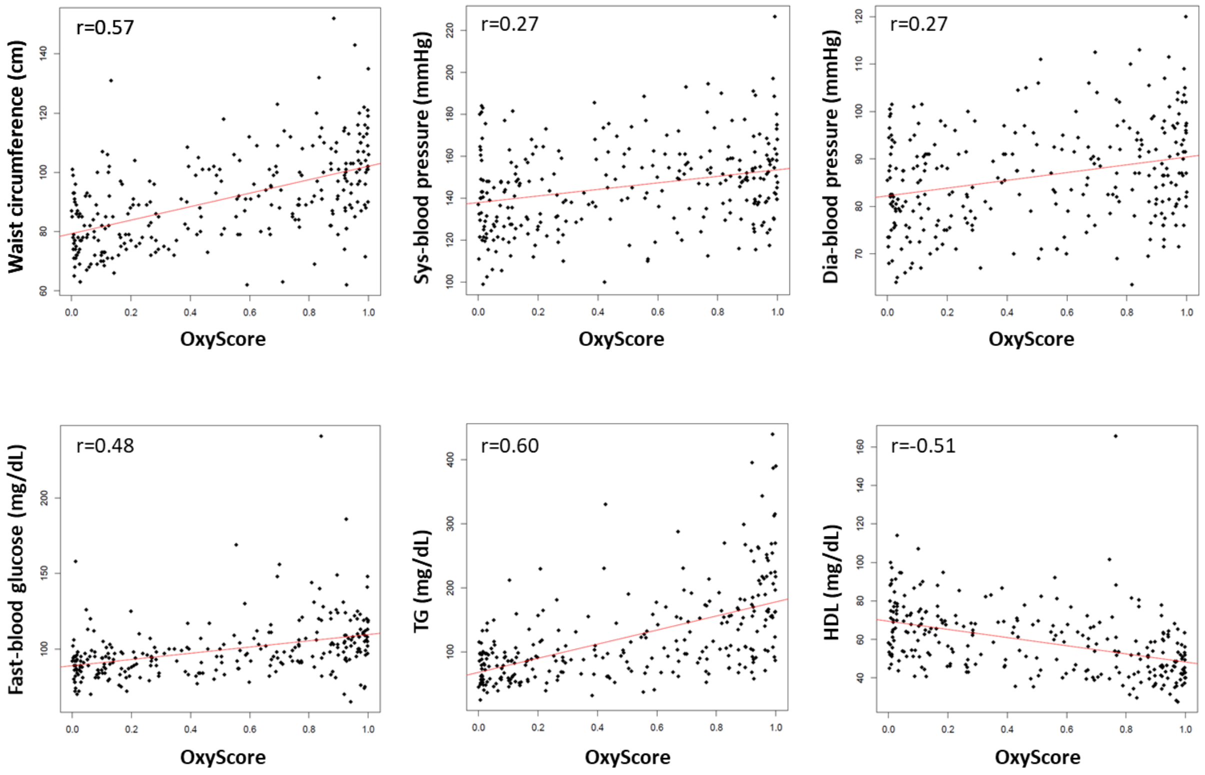

*Supplemental Figure S4. Correlation of the candidate oxylipins with the CardMetS criteria*

Spearman correlations were established in the Discovery study between each candidate oxylipins and the cardiometabolic criteria.

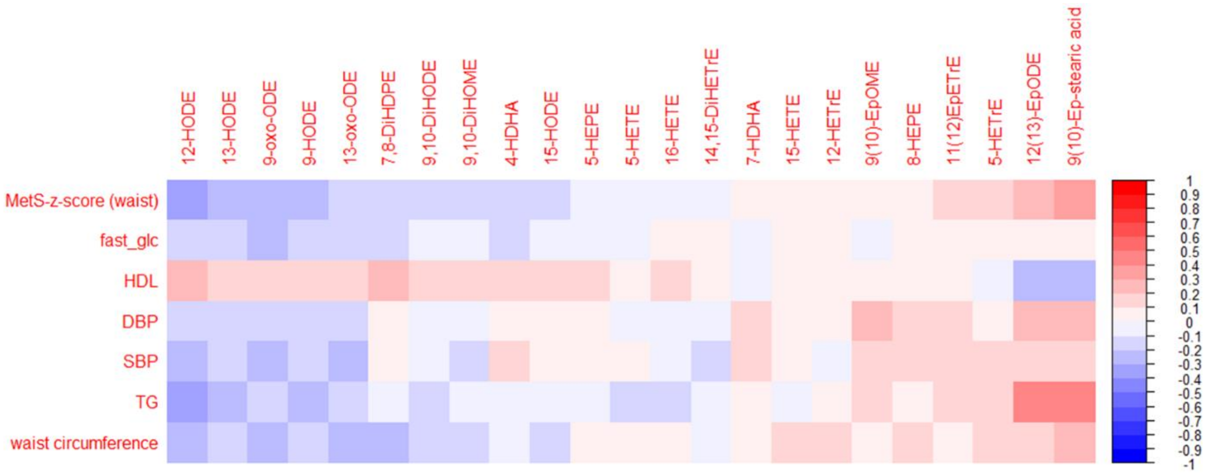

1. Mainka, M., Dalle, C., Pétéra, M., Dalloux-Chioccioli, J., Kampschulte, N., Ostermann, A. I., et al. (2020) Harmonized procedures lead to comparable quantification of total oxylipins across laboratories. J Lipid Res. 61(11):1424-36.
2. Giacomoni, F., Le Corguillé, G., Monsoor, M., Landi, M., Pericard, P., Pétéra, M., et al. (2015) Workflow4Metabolomics: a collaborative research infrastructure for computational metabolomics. Bioinformatics. 31(9):1493-5.
3. van der Kloet, F. M., Bobeldijk, I., Verheij, E. R., Jellema, R. H. (2009) Analytical error reduction using single point calibration for accurate and precise metabolomic phenotyping. J Proteome Res. 8(11):5132-41.
4. Koch, E., Mainka, M., Dalle, C., Ostermann, A. I., Rund, K. M., Kutzner, L., et al. (2020) Stability of oxylipins during plasma generation and long-term storage. Talanta. 217:121074.
