## Supplemental missing data for "Oxylipin profiling identifies a mechanistic signature of metabolic syndrome: results from two independent cohorts"

| Discovery study | 8,9-DiHETrE | 11,12-DiHETrE | 9,10,11-TriHOME | | 9,10-DiHODE | | 17(18)-EpETE | | 10,11-DiHDPE | | 9(10)-EpODE | 12(13)-EpODE | 16-B1-PhytoP | 9-L1-PhytoP | 13-gamma-HOTrE | 17-HETE | 18-HETE | 20-HETE | 12-HHTrE |
|---|---|---|---|---|---|---|---|---|---|---|---|---|---|---|---|---|---|---|---|
| Control (n = 137) (%) | 20 | 23 | 1 | 1 | 4 | 2 | 31 | 3 | 16 | 25 | 7 | 2 | 19 | 4 | | 34 | 2 | 14 | 19 |
| Case (n = 137) (%) | 13 | 18 | 1 | | 3 | | 20 | 7 | 10 | 23 | 2 | | 18 | 4 | 1 | 27 | 2 | 10 | 27 |
| Total (%) | 17 | 20 | 1 | | 4 | | 30 | | 36 | | 5 | 1 | 19 | 4 | 0,4 | 30 | 2 | 12 | 23 |

| Discovery study | 5(S),6(R)-DiHETE (ARA) | 5(S),6(S)-DiHETE (ARA) | 15-F2t-IsoP (8-iso-PGF2a) | 13,14-dihydro-15-oxo-15-F2t-IsoP | 8(9)-EpETE | 11(12)-EpETE | 14(15)-EpETE | 7(S),17(S)-DiH-n3DPA | 4-oxo-DHA | MaR2 | 7,8-DiHDPE | 19,20-DiHDPE | 6-trans-LTB4 | 6-trans-12-epi-LTB4 | 9-HODE | 13-oxo-ODE | 5-HETE | 9-HETE | 8-HEPE |
|---|---|---|---|---|---|---|---|---|---|---|---|---|---|---|---|---|---|---|---|
| Control (n = 137) (%) | 1 | 5 | 26 | 28 | 7 | 21 | | 6 | 30 | 11 | 16 | 1 | 2 | 17 | 37 | 8 | 7 | 49 | 3 |
| Case (n = 137) (%) | 2 | 4 | 29 | 34 | 4 | 15 | 1 | 2 | 30 | 7 | 16 | 1 | 5 | 17 | 26 | 6 | 8 | 42 | 2 |
| Total (%) | 1 | 4 | 28 | 31 | 5 | 18 | 0,4 | 4 | 30 | 9 | 16 | 1 | 4 | 17 | 31 | 7 | 8 | 46 | 3 |

| Replication study | 13-HODE | 9(10)-EpOME | 9-HODE | 13-oxo-ODE | | 9(10)-ep-stearic-acid | | 12(13)-EpOME | | 12-HEPE | 10,11-DiHDPE | 10-HDHA | 11-HDHA | 12(13)-EpODE | 13-oxo-OTrE | 14,15-DiHETrE | 14-HDHA |
|---|---|---|---|---|---|---|---|---|---|---|---|---|---|---|---|---|---|
| Control (n = 101) (%) | 1 | 17 | | 7 | | | 3 | 22 | 58 | 13 | 9 | 2 | 10 | 11 | 68 | 1 | 33 |
| Case (n = 101) (%) | 2 | 17 | 1 | 7 | 1 | 1 | 3 | 21 | 58 | 13 | 11 | | 6 | 13 | 73 | | 29 |
| Total (%) | 1 | 17 | 0 | 7 | | 3 | | 80 | | 13 | 10 | 1 | 8 | 12 | 71 | 0 | 31 |

| Replication study | 15-HEPE | 15-HODE | 16,17-DiHDPE | 16-HETE | 17(18)-EpETE | 17-HDHA | 18-HEPE | 20-HDHA | 5(R,S)-F2t-IsoP | 5-HEPE | 7(8)-EpDPE | 7-HDHA | 8(9)-EpETrE | 8-HDHA | 8-HEPE | 9,10,11-TriHOME | 9,10-DiHODE | NPDx-1 | tetranor-12(S)-HETE |
|---|---|---|---|---|---|---|---|---|---|---|---|---|---|---|---|---|---|---|---|
| Control (n = 101) (%) | 50 | 2 | 13 | 26 | 6 | 58 | 89 | 3 | 82 | 3 | 1 | 4 | 16 | 3 | 16 | 18 | 6 | 92 | 75 |
| Case (n = 101) (%) | 51 | 2 | 18 | 24 | 4 | 63 | 85 | 5 | 72 | | 1 | 3 | 17 | 3 | 14 | 24 | 13 | 90 | 57 |
| Total (%) | 50 | 2 | 15 | 25 | 5 | 61 | 87 | 4 | 76 | 1 | 1 | 3 | 16 | 3 | 15 | 21 | 9 | 91 | 66 |

| Replication study | 12(S)-HETrE | | 14(15)-EpETE | | 5(S)-HETrE | | 7,8-DiHDPE | | 8(9)-EpETE | | 8(S)-HETrE | | 9-HOTrE | | 11-HEPE | 13-HOTrE | 15(S)-HETrE | 15-HETE | 19,20-DiHDPE | 9(10)-EpODE | 9-HEPE | 9-HETE |
|---|---|---|---|---|---|---|---|---|---|---|---|---|---|---|---|---|---|---|---|---|---|---|
| Control (n = 101) (%) | 17 | 14 | 4 | | 9 | 2 | 13 | | 29 | 1 | 36 | 32 | 20 | 28 | 97 | 38 | 3 | 7 | 30 | 29 | 93 | 29 |
| Case (n = 101) (%) | 17 | 13 | 2 | 1 | 2 | | 21 | 1 | 27 | | 28 | 31 | 17 | 26 | 99 | 38 | | 4 | 31 | 29 | 95 | 29 |
| Total (%) | 24 | | 3 | | 6 | | 17 | | 28 | | 45 | | 37 | | 98 | 38 | 1 | 5 | 30 | 29 | 94 | 29 |

| <LLOQ (flag 1) | >ULOQ (flag 2) | N.A. (flag 6) | No integration possible (flag 7) | % > 30% |
|---|---|---|---|---|

Table 3. Pourcentage of missing data for each studies, according to the different flag
